# Optimising health and economic impacts of COVID-19 vaccine prioritisation strategies in the WHO European Region

**DOI:** 10.1101/2021.07.09.21260272

**Authors:** Yang Liu, Frank G. Sandmann, Rosanna C. Barnard, Carl A.B. Pearson, CMMID COVID-19 Working Group, Roberta Pastore, Richard Pebody, Stefan Flasche, Mark Jit

## Abstract

**Background:** Countries in the World Health Organization (WHO) European Region differ in terms of the COVID-19 vaccine roll-out speed. We evaluated the health and economic impact of different age-based vaccine prioritisation strategies across this demographically and socio-economically diverse region.

**Methods:** We fitted country-specific age-stratified compartmental transmission models to reported COVID-19 mortality in the WHO European Region to inform the immunity level before vaccine roll-out. Building upon broad recommendations from the WHO Strategic Advisory Group of Experts on Immunisation (SAGE), we examined four strategies that prioritise: all adults (V+), younger (20-59 year-olds) followed by older adults (60+) (V20), older followed by younger adults (V60), and the oldest adults (75+) (V75) followed by incremental expansion to successively younger five-year age groups. We explored four roll-out scenarios based on projections or recent observations (R1-4) - the slowest scenario (R1) covers 30% of the total population by December 2022 and the fastest (R4) 80% by December 2021. Five decision-making metrics were summarised over 2021-22: mortality, morbidity, and losses in comorbidity-adjusted life expectancy (cLE), comorbidity- and quality-adjusted life years (cQALY), and the value of human capital (HC). Six sets of infection-blocking and disease-reducing vaccine efficacies were considered.

**Findings:** The optimal age-based vaccine prioritisation strategies were sensitive to country characteristics, decision-making metrics and roll-out speeds. Overall, V60 consistently performed better than or comparably to V75. There were greater benefits in prioritising older adults when roll-out is slow and when VE is low. Under faster roll-out, V+ was the most desirable option.

**Interpretation:** A prioritisation strategy involving more age-based stages (V75) does not necessarily lead to better health and economic outcomes than targeting broad age groups (V60). Countries expecting a slow vaccine roll-out may particularly benefit from prioritising older adults.

**Funding:** World Health Organization, Bill and Melinda Gates Foundation, the Medical Research Council (United Kingdom), the National Institute of Health Research (United Kingdom), the European Commission, the Foreign, Commonwealth and Development Office (United Kingdom), Wellcome Trust

**Research in Context:** *Evidence before this study:* We searched PubMed and medRxiv for articles published in English from inception to 9 Jun 2021, with the search terms: (“COVID-19” OR “SARS-CoV-2”) AND (“priorit*) AND (“model*”) AND (“vaccin*”) and identified 66 studies on vaccine prioritization strategies. Of the 25 studies that compared two or more age-based prioritisation strategies, 12 found that targeting younger adults minimised infections while targeting older adults minimised mortality; an additional handful of studies found similar outcomes between different age-based prioritisation strategies where large outbreaks had already occurred. However, only two studies have explored age-based vaccine prioritisation using models calibrated to observed outbreaks in more than one country, and no study has explored the effectiveness of vaccine prioritisation strategies across settings with different population structures, contact patterns, and outbreak history.

*Added-value of this study:* We evaluated various age-based vaccine prioritisation strategies for 38 countries in the WHO European Region using various health and economic outcomes for decision-making, by parameterising models using observed outbreak history, known epidemiologic and vaccine characteristics, and a range of realistic vaccine roll-out scenarios. We showed that while targeting older adults was generally advantageous, broadly targeting everyone above 60 years might perform better than or comparably to a more detailed strategy that targeted the oldest age group above 75 years followed by those in the next younger five-year age band. Rapid vaccine roll-out has only been observed in a small number of countries. If vaccine coverage can reach 80% by the end of 2021, prioritising older adults may not be optimal in terms of health and economic impact. Lower vaccine efficacy was associated with greater relative benefits only under relatively slow roll-out scenarios considered.

*Implication of all the available evidence:* COVID-19 vaccine prioritization strategies that require more precise targeting of individuals of a specific and narrow age range may not necessarily lead to better outcomes compared to strategies that prioritise populations across broader age ranges. In the WHO European Region, prioritising all adults equally or younger adults first will only optimise health and economic impact when roll-out is rapid, which may raise between-country equity issues given the global demand for COVID-19 vaccines.

## Introduction

The COVID-19 pandemic poses unprecedented challenges to public health, health systems and economies globally. Extraordinary efforts and resources have been committed to the development and roll-out of COVID-19 vaccines and vaccination programmes.^1^ These global efforts have led to successful vaccine development at an unprecedented speed.

While some countries have signed bilateral advanced purchasing agreements with vaccine manufacturers to independently procure enough vaccine doses to cover large proportions of their populations, many low- and middle-income countries (LMICs) do not have the resources for such an option.^2^ Globally coordinated efforts to roll out COVID-19 vaccines are therefore required to achieve equitable vaccine distribution and to control the COVID-19 pandemic.

The global initiative “COVID-19 Vaccines Global Access” (COVAX) has been set up to ensure equitable vaccine access across countries.^3^ However, the speed at which vaccines become available through COVAX is constrained by production and logistical capacities. In the interim vaccine distribution forecast published in February 2021, COVAX was projected to deliver vaccines to cover approximately 3% of the total population in the 145 COVAX facility participant countries by mid-2021, and up to 20% of those populations by the end of 2021.^3,4^ Among the 53 countries in the WHO European Region, 16 Member States may follow this projection as non-Advanced Market Commitment (AMC) donors.^3^

Given the diverse supply conditions, countries in WHO European Region have decided on different prioritisation strategies for vaccinating against COVID-19. Besides prioritising healthcare workers based on elevated exposure risks, countries have also been recommended to implement age-based vaccine prioritisation strategies targeting older adults.^5,6^ However, the specific age cut-off for “older adults” have not been consistent, ranging from 50 to 80 years within the region.^7^ Some countries have not set out to prioritise by age at all.^7^ Elsewhere in the world, some countries have chosen to prioritise younger adults instead. The overall impacts of different vaccine prioritisation strategies under diverse supply conditions is, therefore, an instrumental piece of evidence supporting COVID-19 vaccine policies.

This study evaluates different age-based vaccine prioritisation strategies given different vaccine supply conditions between 2021 and 2022 in the WHO European Region. We identify strategies that maximise the health and economic impacts of COVID-19 vaccines, measured by five decision-making metrics which allowed us to explore the trade-offs between minimising COVID-19 mortality and morbidity. We consider the known epidemiology of SARS-CoV-2, demographic factors, government COVID-19 response policy stringency, human mobility in each country, and a wide range of hypotheses about vaccine mechanisms and characteristics.

## Methods

### Model Framework

Our model framework consisted of two stages (Figure 1). The fitting stage was to estimate the proportion of each age group in every country who were no longer susceptible to SARS-CoV-2 by 01 Jan 2021. The projection stage relied on the results from the fitting stage, vaccine-related assumptions, health and economic impact parameters, and projected mobility changes related to expected public health and social measures. Using these, we estimated the health and economic outcomes for each country under different vaccine supply conditions (expressed as roll-out scenarios) and prioritisation strategies by December 2022.

**Figure 1.**
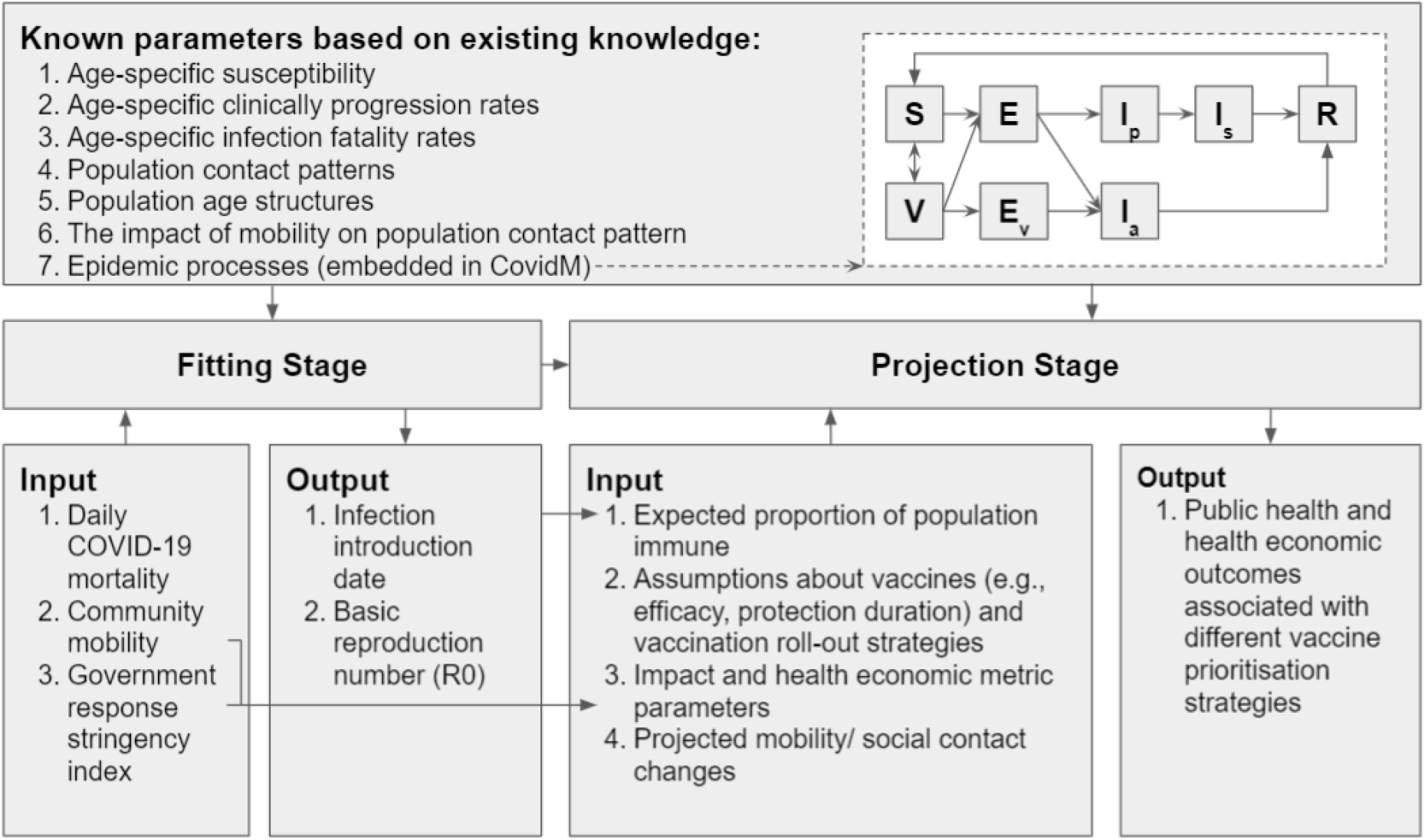
Model Framework. This figure describes the overall model framework of the study, which consists of the fitting and projection stages. The “known parameters based on existing knowledge” were used in both fitting and projection stages. The remaining input was used in only one of the stages as specified. In the conceptual diagram of the model: S - Susceptible; E - Exposed; V - Vaccinated; E_v_ - Exposed among vaccinated; I_p_ - presymptomatic & infectious; I_s_ - symptomatic & infectious; I_a_ - asymptomatic & infectious; R - Removed.

For both stages, we used a previously described COVID-19 compartmental dynamic transmission model, CovidM.^8^ This model includes 16 age groups defined by five-year age bands (from 0-4 to 75+), incorporates differences in infectiousness among pre-symptomatic, asymptomatic, and symptomatic infections; age-dependent clinical fractions (e.g. the proportion of infected cases that are symptomatic);^9^ age-dependent susceptibilities;^9^ and age-dependent infection-fatality ratios.^10^ We used country-specific population age-structures^11^ and age-stratified contacts rates^12^ to capture population dynamics (Figure 2, A and B). A complete table summarising input variables and assumptions is presented in Appendix 1.

**Figure 2.**
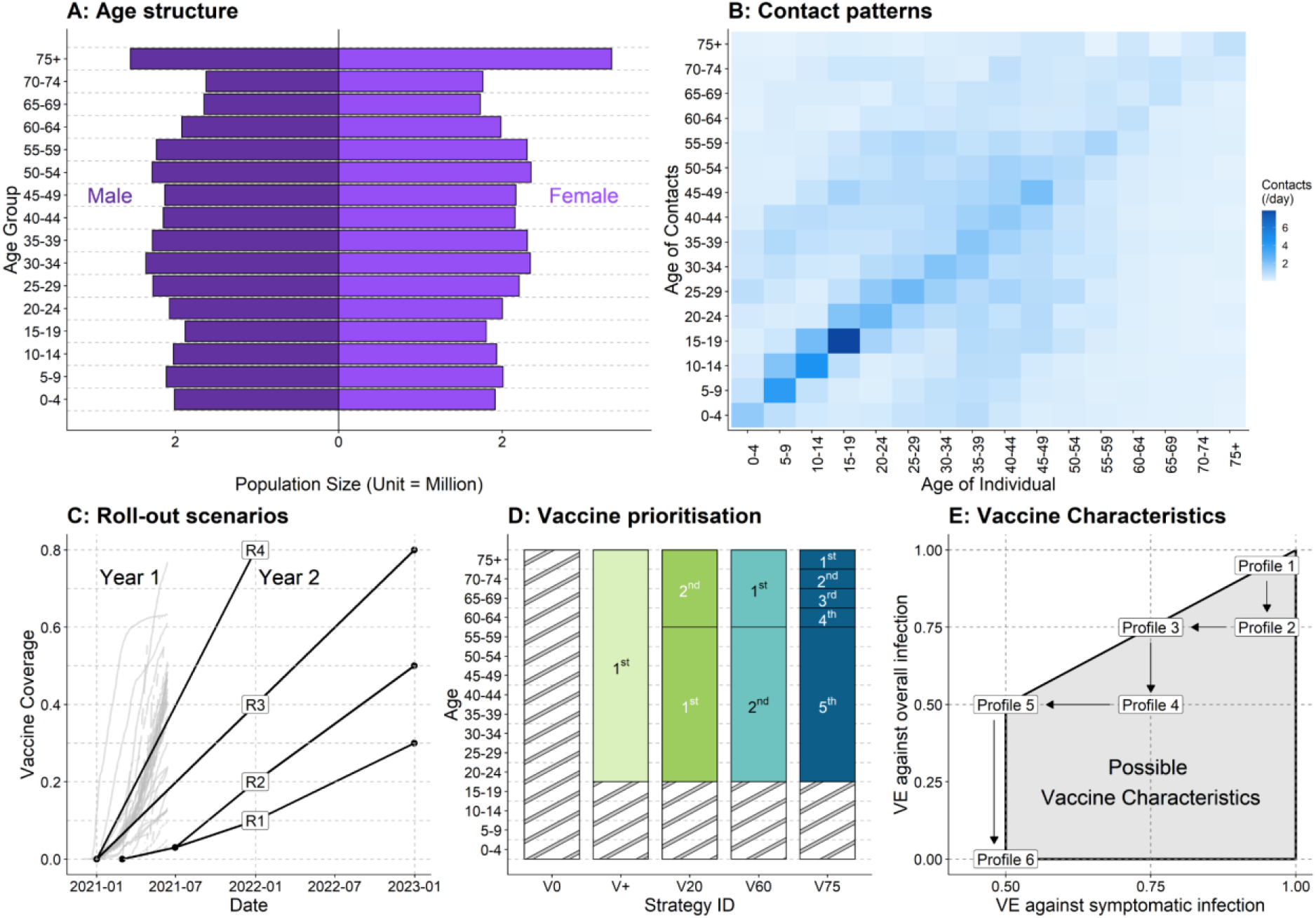
Key inputs and assumptions. (A) Example population age structure (for the United Kingdom, unit = million).^11^ (B) Example age-specific within-population contact patterns (for the United Kingdom).^12^ (C) Vaccine roll-out scenarios and the respective proportions of populations expected to be covered at different time points. Note that under different vaccine roll-out scenarios, the starting time of vaccination programs may diff. Grey lines in the background represent observed country-level vaccine uptakes (of the first dose) over time reported in the WHO European Region.^19^ (D) Vaccine prioritisation strategies. Hatched areas indicate when no vaccine is allocated into the corresponding age groups. (E) Vaccine profiles consisting of vaccine efficacy against infection and disease.

### Fitting stage

Country-specific infection introduction dates and basic reproduction numbers (R0s) were obtained by fitting the model deterministically to reported COVID-19 mortality using maximum likelihood estimation (MLE) and differential evolution global optimization algorithms assuming a Poisson likelihood function for daily mortality.^13^ The infection introduction date was constrained to a 90-day window before the first reported COVID-19 death in each country. The R0s were constrained to the 1.5 to 5 range. We also generated 100 stochastic realisations of mortality over time to evaluate the overall performance of the fitting stage.

We used smoothed country-specific daily reported COVID-19 mortality (i.e. right-aligned seven-day moving averages) based on raw data from the 2019 Novel Coronavirus Visual Dashboard, most of which has been collected from the government reports.^14^ Age-dependent social contact patterns were captured by country-specific synthetic contact matrices, which include information from different contact settings: *work, school, home*, and *others*.^12^ However, social contacts have deviated substantially from their pre-pandemic levels. Thus, we used Google mobility data to approximate the changes in contact patterns in *work* and *other* settings during the pandemic.^15,16^ Additionally, social contacts in the *school* setting were based on school-related non-pharmaceutical intervention (NPI) stringency.^17^ Contacts at *home* were assumed to stay unchanged compared to pre-pandemic periods. Please see Appendix 2 for more details.

Evidence suggests COVID-19 mortality may have been higher than reported in many countries.^18^ To account for this possibility, we repeated the fitting stage estimating for an additional parameter - a country-specific temporally-invariant mortality underreporting rate, as a sensitivity analysis.

### Projection stage

We assume that social contact patterns (*work* and *other*) gradually recover over a year after March 2021 but never fully return to pre-pandemic levels, due to long-term policies and behavioural changes. Social contacts in the *school* setting followed school terms. More details are presented in Appendix 2.

We examined four roll-out scenarios: R1, R2, R3 and R4 (Figure 2, C). R2 resembles the projected roll-out projections under COVAX.^3,4^ In R1, vaccine roll-out is slower and final uptake lower compared to R2. Contributors may include production delay, logistical challenges in delivering vaccines, or vaccine demand (e.g. vaccine confidence). R1 and R2 both start from 01 Mar 2021. R4 resembles the trajectories of the rapid adopters of the region.^19^ R3 achieves the same coverage as R4, but at a slower pace. R3 and R4 both start from 01 Jan 2021. In the WHO European Region, as of July 2021, 29 countries have rolled out vaccination at an equal or greater rate than R4, 8 countries at a rate between R3 and R4, 11 countries at a rate between R1/2 and R3, and the remaining five countries have not reported vaccine uptake yet.^19^

We examined four vaccine prioritisation strategies in addition to the counterfactual where no vaccines were delivered (V0) (Figure 2, D). In V+, the entire population aged 20+ years received vaccines simultaneously. In strategy V20, younger adults (20-59 year-olds) received vaccines before older adults (60+ year-olds). In both V60 and V75, older adults (60+ year-olds) were vaccinated first; however, in V60 they were all vaccinated simultaneously, whereas in V75 age groups were prioritised in 5-year age bands, starting from those above 75 years of age. In this study, the vaccine roll-out of an age group was considered complete when 90% of that age group had received a vaccine for 60+ years old and 70% for 20-59 year-olds. These targets are consistent with optimistic roll-out objectives and intended vaccine uptake observed in the WHO European Region.^1,20^ We additionally explored the condition when adolescents (10-19 year-olds) become eligible for the vaccines.

There is still considerable uncertainty around the immunity against SARS-CoV-2. In this study, we assumed that vaccinees were protected 14 days following receipt of a vaccine dose.^21^ Based on a recent cohort study,^22^ we assumed that protection gained from natural SARS-CoV-2 infection would wane exponentially with a mean duration of 3 years. For vaccine-induced protection, we assumed it would wane exponentially with a mean duration of 52 weeks (with 3 years as sensitivity analysis). Infection-blocking and disease-reducing mechanisms of vaccines were both modelled. As baseline vaccine efficacy (VE), we assume that vaccination prevents 95% of infections and diseases based on the best-performing vaccines in clinical trials and post-introduction observational data.^21,23^ Additionally, we explored a range of vaccine profiles (see Figure 2, E). The worst performing vaccine we included prevented 50% of diseases and had no effect on infection.

We used an ordinal logistic regression model to explore if the optimal vaccine prioritisation strategies identified are associated with country-specific characteristics such as population sizes, age structures, contact patterns, proportions of non-susceptible individuals by 01 Jan 2021, and roll-out scenarios.

### Outcomes

We assessed the impact of different vaccine prioritisation strategies in each country on five health and economic decision-making metrics summarised over 01 Jan 2021∼31 Dec 2022: (1) mortality, (2) cases, (3) comorbidity-adjusted life expectancy (cLE) loss, (4) comorbidity- and quality-adjusted life years (cQALY) loss, and (5) value of human capital (HC) loss. The optimal strategy minimises these outcomes. Since the top-ranking strategies may have similar effects, we additionally calculated the regional totals for each health and economic metric when one strategy is applied to the entire region. We briefly define (3)-(5) below and provide more details in Appendix 3.

Comorbidity-adjusted life expectancy (cLE) is a measure of the number of years an individual would be expected to live had they not died prematurely, based on the life expectancy of an individual with an average level of comorbidities.^24^ These values are country- and age-specific.

Comorbidity- and quality-adjusted life years (cQALYs) are similar to cLE, but weigh the years someone is alive based on their health-related quality of life.^25^ The cQALY loss due to premature COVID-19 mortality was calculated using the average age-dependent population norms available for seven European countries and country-specific life expectancies, assuming the QALY norms to be the same in other countries in the WHO European Region.^26^ We estimated that each symptomatic COVID-19 case leads to the loss of 0.0307 QALYs (see Appendix 1), which incorporates detriments due to non-hospitalised illness episodes, hospitalisations, intensive-care treatment, and post-acute symptoms (long COVID). Short-term vaccine side effects were assumed to occur at 50% probability and to lead to a loss of 1 quality-adjusted life day per vaccinee.^27,28^

The human capital (HC) method weighs the number of life-years someone loses by the country-specific GDP per capita, as a proxy for the value of that individual’s lost lifetime productivity. These values are not adjusted for age or sex, since there is growing consensus that doing so undervalues unpaid labour.^29^ We used the World Bank’s country-level GDP per capita estimates.^30^

The analyses were conducted using R (4.0.4).^31^ An R Shiny application is available at https://cmmid-lshtm.shinyapps.io/demo/ to enable users to explore additional sets of parameters not covered in this study.

## Results

### Model fitting

Of the 53 WHO European Region Member States, we were able to fit our model to daily reported COVID-19 mortality for 38. The remaining 15 either reported no COVID-19 (n = 1), reported too few COVID-19 deaths (<10 deaths/day) to allow reliable inference (n = 12), or had reported major changes to case definitions (n = 2), and were thus excluded from further analyses. The outcomes of the fitting process are presented in Figure 3, A-C, using Georgia, Hungary and the United Kingdom as examples. Results for all other countries have been archived in our Zenodo repository.^32^

**Figure 3.**
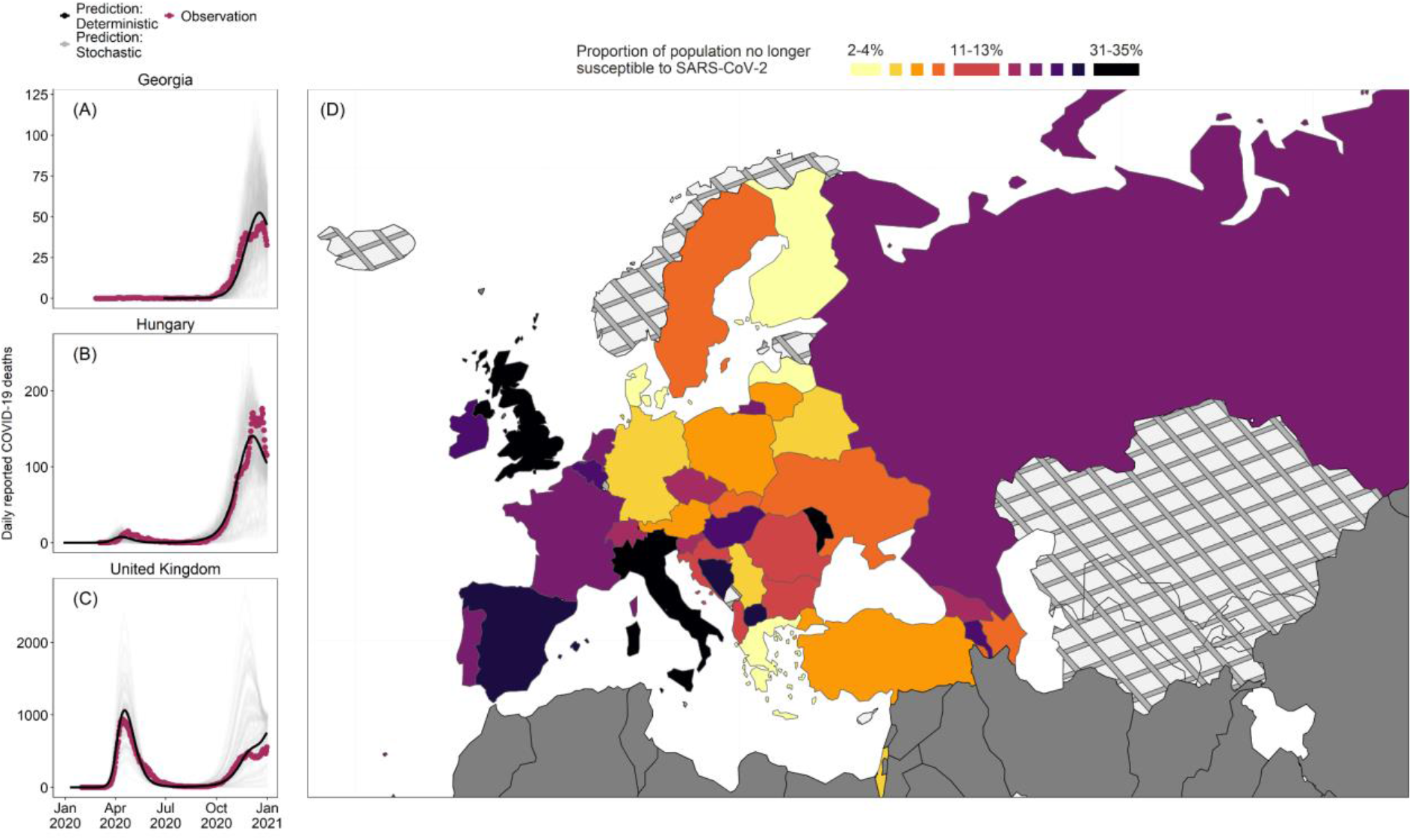
Results of the fitting stage. (A-C) Comparisons between observed COVID-19 (purple line), predicted COVID-19 deaths using a deterministic realisation using fitted parameters (black line), and 100 stochastic realisations using the same fitted parameters (grey lines) in Georgia, Hungary, and the United Kingdom. (D) The estimated proportions of individuals no longer susceptible (infectious or recovered) to SARS-CoV-2 infection on 01 Jan 2021 by deciles. Countries marked by crosshatch patterns are those that were not included in the fitting stage; countries marked by solid grey are outside the WHO European Region. Shapefiles were downloaded from Eurostat GISCO.^34^

We found that the inferred infection introduction date in most countries occurred in January 2020 (n =38, IQR: 25 Dec 2019, 14 Jan 2020) (Appendix 4). The median R0 estimates was 1.67 (n = 38, IQR: 1.58, 1.85) (Appendix 4). These estimates are relatively low compared to existing literature,^33^ possibly due to NPIs that are not fully captured by the intensity of mobility and social contacts (e.g. mask-wearing). Using these inferred parameters, we estimated the median proportion of individuals no longer susceptible to SARS-CoV-2 by 01 Jan 2021 to be 13.09% (n = 38, IQR: 6.94%, 21.63%) (Figure 3, D).

### Optimal prioritisation strategies

We observed substantial differences in the optimal vaccine prioritisation strategy by decision-making metrics. Under R1 and R2, the optimal strategy is V75 to minimise COVID-19 deaths and V20 to minimise COVID-19 cases (Figure 4 A-B, F-G). Under R1 and based on cLE, cQALY or HC losses, most countries had V60 as their optimal strategy although overall the performance of V+, V20 and V60 was similar when applied to the entire region, leading to 3-7% more losses compared to when local optimal strategies were used (Figure 4, C-E). Under R2, although there were a comparable number of countries with V+, V20 and V60 as their optimal strategy based on cLE, cQALY and HC losses, V+ was the best performing strategy when applied to the entire region (Figure 4 H-J, Appendix 5).

**Figure 4.**
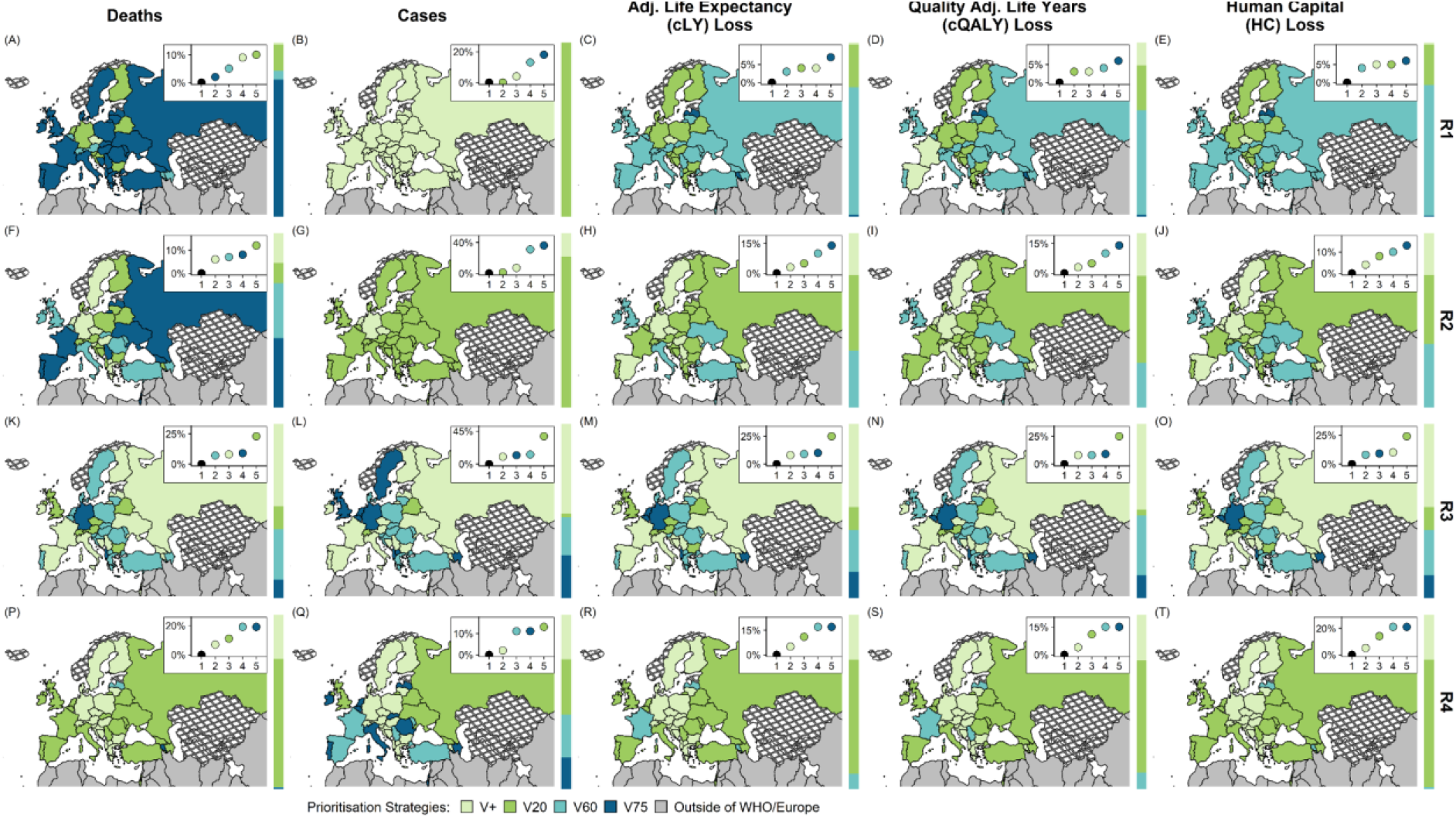
Optimal vaccine prioritisation strategies under different roll-out scenarios and decision-making metrics. Main panel - Optimal strategies across the WHO European Region that minimise COVID-19 deaths, cases, losses in adjusted life expectancy (cLE), quality-adjusted life-years (cQALY), and human capital (HC) as decision-making metrics. Top right insert within each panel - y-axis: Difference in outcome (totalled over the region) when a given prioritisation strategy is used across the entire WHO European Region compared to if the optimal prioritisation strategy in each country is used (black) x-axis: ranking. Sidebars to the right of each panel - the proportion of total population for which each prioritisation strategy is optimal. Shapefiles are downloaded from Eurostat GISCO;^34^ countries marked by crosshatch patterns are those that were not included in the projection stage.

Under R3, V+, V60 and V75 have roughly equal performance when applied to the entire region regardless of metric (Figure 4 K-O), leading to an 8-13% increase compared to when the optimal strategy for each country was used instead using all metrics. However, under R4, not prioritising by age (V+) becomes the optimal strategy in most countries (Figure 4, P-T, Appendix 5). The comparative advantage of V+ under R4 is robust to a longer assumed duration of vaccine-induced immunity (i.e. 3 years) (Appendix 6).

The results of the ordinal logistic regression show that the only factors that were significantly associated with the optimal vaccine prioritisation strategies (with *p*-values < 0.05) were roll-out scenarios (Appendix 7). Faster roll-out scenarios were associated with having optimal strategies that target younger or all adults. Additionally, there was weak evidence supporting the association between the intensity of adult to older adult contacts with having optimal strategies that prioritise older adults (*p-* value < 0.1).

In our sensitivity analysis that incorporated underreporting of COVID-19 mortality, we estimated higher proportions of non-susceptible individuals than baseline scenarios. In these cases, the benefits of targeting older adults diminished under R3 while other results remained broadly unchanged (Appendix 8).

We noticed that the optimal prioritisation strategies can be sensitive to the decision time frame considered after summarising decision metrics over 6, 12, and 18 months following the start of vaccination. Although results are robust under R1 and R2, we found substantial variability in optimal strategies across decision-making metrics under R3 and R4 (Appendix 9). The temporal cut-off presented as the baseline scenario in this study (i.e. drawing a deadline at 2022) was decided through discussion with stakeholders. Vaccines may delay upcoming epidemic peaks, and having a temporal cut-off may favour later outbreaks over sooner despite epidemic sizes, leading to biased interpretations. Lastly, our model shows that vaccinating adolescents with younger adults may bring additional health and economic benefits (Appendix 10).

### Sensitivity analysis by vaccine characteristics

In Figure 5, we present the results under R2 and R3 (additional results under R1 and R4 are presented in Appendix 11). The optimal vaccine prioritisation strategy is sensitive to the vaccine profile assumed. Under R2, a vaccine product with lower VEs (i.e. profiles 5 and 6) would result in prioritising older adults (i.e. V60) being the optimal strategy in many countries in terms of minimising deaths, and losses in cLE, cQALY and HC. Under R3 the differences between V+, V60 and V75 are small. The optimal strategy varies widely across countries when VE changes. When VE is low (profile 5 and 6), slightly more countries had V20, and fewer countries had V+ as their optimal strategy compared to when VE is high (profile 1 and 2).

**Figure 5.**
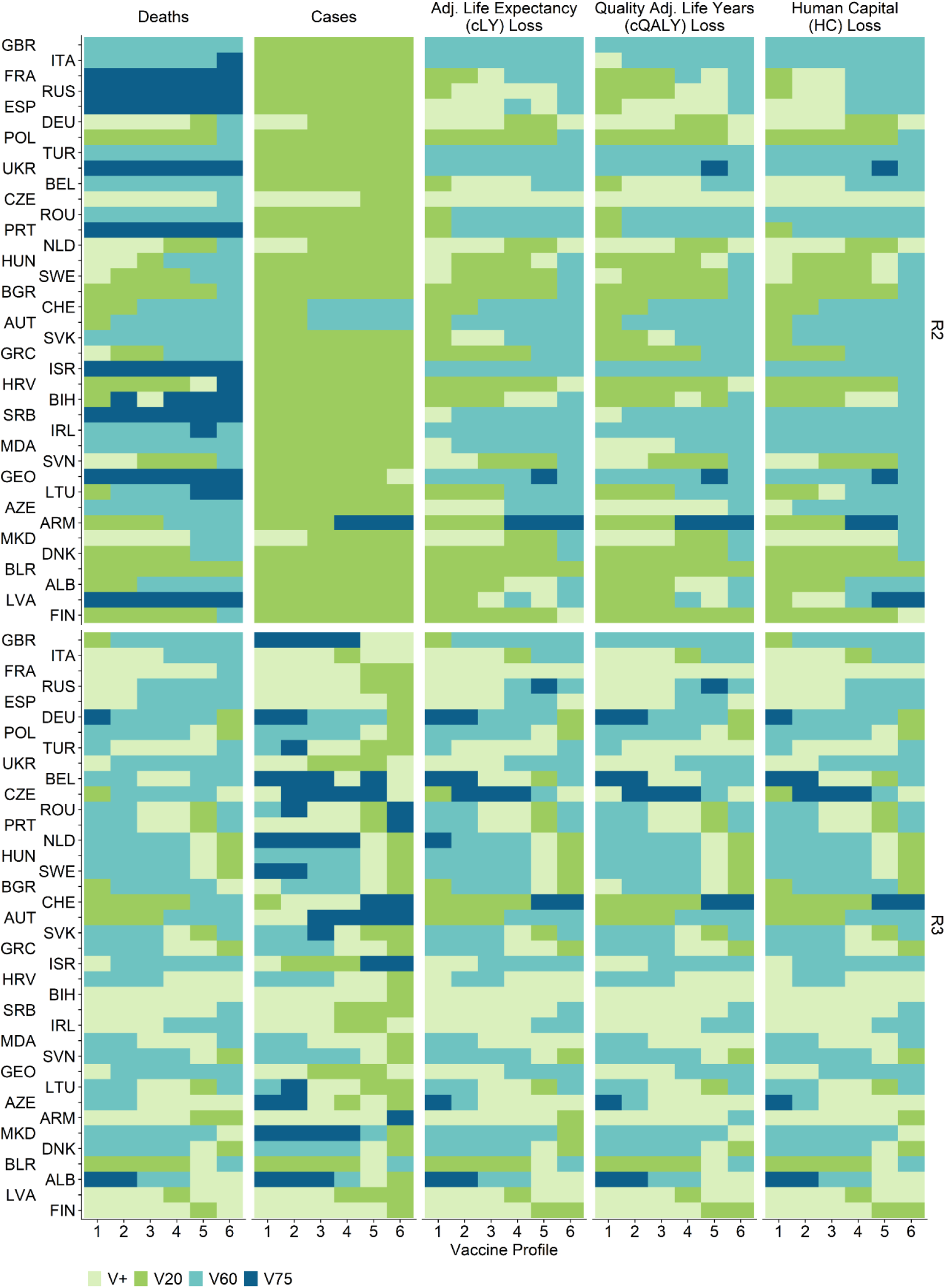
Optimal vaccine prioritisation strategies given different vaccine profiles under R2 and R3. Optimal strategy for each country and vaccine profile while minimising mortality, morbidity, and losses in adjusted life expectancy (cLE), quality-adjusted life-years (cQALY), and human capital (HC) for 38 countries in the WHO European Region. Countries are arranged in the order of the estimated proportions of the population no longer susceptible to SARS-CoV-2 on 01 Jan 2021 (descending). Countries are labelled by their World Bank country codes - a reference table can be found in Appendix 1.

## Discussion

Using a transmission model of SARS-CoV-2 fitted to reported COVID-19 mortality data in the WHO European Region, this study evaluates different vaccine prioritisation strategies under a range of possible vaccine roll-out scenarios. We found that age-based vaccine prioritisation strategies with more stages do not necessarily perform better. A two-stage approach that prioritises all older adults (V60) consistently performs better than or comparably to a five-stage approach that initially prioritises the oldest age group and only extends to the next five-year age group younger when vaccination goals of the last group have been met (V75). Depending on local context, the choice between V60 and V75 could depend on other factors beyond health and economic impacts (e.g. ease of roll-out, health system characteristics).

We found that when vaccine supply and delivery capacity are in place to reach 80% coverage by the end of 2021, the benefits of precisely targeting older adults diminish. Prioritising older adults may be more valuable to countries that expect a longer timeline for vaccine uptake to increase (e.g. nearly half of the WHO European Region Member States). These findings raise issues around within-country and between-country health equity. Countries with sufficient supplies of vaccines may find that they maximise benefits domestically by offering vaccination without any age restrictions, as this could expedite roll-out. However, there are insufficient supplies of COVID-19 vaccines globally to allow all countries to pursue such a strategy. Increased supplies for one country come at the expense of another.^35^

Evidence on infection-blocking and disease-reducing VE of different vaccine products is emerging. Here, we examined six different vaccine profiles, ranging from a highly effective vaccine that prevents both infection and disease at 95%, to a vaccine that only prevents 50% of disease and does not provide any protection against infection. This range is designed around the vaccine products currently licensed in the WHO European Region. If vaccines become disease-reducing but not infection-blocking, more countries expecting slower roll-out may prioritise older adults (V60 or V75) given that the severity of disease burden increases by age.

### Strengths and limitations

While existing literature focuses on evaluating COVID-19 vaccine prioritisation in any particular city or country, our study is the first to investigate the impact of such strategies under different circumstances across a large region. We demonstrated that different age structures, contact patterns, and sizes of existing epidemics may affect the health and economic impacts of vaccination prioritisation strategies. Second, we built our analyses around realistic vaccine roll-out scenarios based on projecting the broad patterns in vaccine roll-out rates across the region. Third, we evaluated a wide range of age-prioritisation strategies, not only under different roll-out scenarios but also at different supply levels (e.g. 3%, 20%, 50% or 80% coverage). The results inform policy recommendations, providing the necessary details for continuous implementation. The models and analyses presented here have benefited from continuous input since late 2020 from the WHO Regional Office for Europe and technical advisors from many countries in the region. Fourth, we included additional decision-making metrics beyond mortality and morbidity, providing a more comprehensive assessment of the various trade-offs that need to be considered between outcomes.^36^ Fifth, we considered various vaccine profiles to account for uncertainty around vaccine characteristics. Last but not the least, new evidence about vaccine action, changes in vaccine supply constraints and the emergence of additional variants of concern (VOCs) may necessitate re-evaluation. To facilitate this, we have developed an online tool to help inform such policy decisions.

We did not examine exposure risk-(e.g. highly connected individuals) or occupation-based (e.g., healthcare providers) vaccine prioritisation strategies. The evaluation of these strategies requires parameters governing their transmission risk (e.g. contact matrices by occupation, coverage and effectiveness of personal protective equipment), most of which are not currently available. We did not project healthcare system outcomes such as hospital bed and intensive care unit occupancy as there are substantial variations across countries and subnational regions in patient pathways, healthcare capacity and hospital organisation.

The fitting stage is based on mortality data before 2021, before the start of vaccine roll-out in some countries and the large-scale emergence of VOCs that are more transmissible, more clinically severe, and potentially immune escaping.^37^ Future research would require vaccine product-specific uptake levels, country-specific underreporting rates for cases (which is far more variable temporally and across countries than mortality underreporting), and VOC-specific surveillance data, which are not easily available across the WHO European Region.

The fitting stage includes a country-specific temporally-invariant underreporting rate of COVID-19 mortality as a part of the sensitivity analysis. This rate may be time-varying;^38^ however incorporating time dependencies would likely make parameter identification difficult. The projection stage uses constant vaccine efficacy values before Dec 2022, which may not remain valid as VOCs spread, although we used a range of vaccine profiles that likely cover performance against the most common variants in the WHO European Region as of mid-2021.^39^ Additionally, we technically modelled the vaccine in use as a single-dose product with a 14-day delay between vaccination and effective protection. These assumptions may be replaced by new evidence on VE and the immunity development process as they emerge.

### Conclusion

In conclusion, we identified optimal age-based vaccine prioritisation strategies under different vaccine roll-out scenarios and vaccine effectiveness profiles at the country level in the WHO European Region. We showed that the benefits of prioritising older adults were more evident for relatively slow vaccine roll-out scenarios. When prioritising older adults, broadly targeting everyone above 60 years consistently performed comparably or better than targeting the oldest adults first followed by the next younger five-year age group - the additional stages does not lead to health or economic gains. Prioritising younger adults or not prioritising by age is only beneficial when vaccines can be rolled out quickly (e.g. reaching 80% of the population vaccinated in 1 year).

## Supporting information

Technical Appendix

## Data Availability

We used publicly available data in this study, cited in the reference list or in the appendix. The CovidM modelling framework used has been published previously and is available on the CMMID COVID-19 GtiHub page. All code used and country-specific intermediate results have been archived via zenodo. An R shiny application is available at https://cmmid-lshtm.shinyapps.io/demo/ to enable users to explore additional sets of parameters not covered in this study.

## Acknowledgements

We graciously thank the following agencies for their support to this work: World Health Organization (202604060), Bill & Melinda Gates Foundation (INV-003174, OPP1184344), European Commission (101003688), Medical Research Council (MC_PC_19065), National Institute of Health Research (200929), Foreign, Commonwealth and Development Office (UK)/ Wellcome Trust (221303/Z/20/Z), Wellcome Trust (208812/Z/17/Z). FGS and MJ were supported by the NIHR Health Protection Research Unit (HPRU) in Modelling and Health Economics, a partnership between Public Health England (PHE), Imperial College London, and the London School of Hygiene & Tropical Medicine (LSHTM; grant code NIHR200908). MJ was supported by the NIHR HPRU in Immunisation at LSHTM in partnership with PHE (grant reference code NIHR200929). We are grateful for the support, comments, and feedback from the members and organizers of WHO’s Regional Working Group on COVID-19 vaccination and development in the European Region, Focus Group 2 “Immunization Strategy and decision making.” We thank Dr Nicholas G Davies for his valuable feedback and for his work on CovidM - much of what this analysis has been built upon.

The following funding sources are acknowledged as providing funding for the working group authors. This research was partly funded by the Bill & Melinda Gates Foundation (INV-001754: MQ; INV-003174: KP; INV-016832: SRP; NTD Modelling Consortium OPP1184344: GFM; OPP1139859: BJQ; OPP1191821: KO’R). BMGF (INV-016832; OPP1157270: KA). CADDE MR/S0195/1 & FAPESP 18/14389-0 (PM). EDCTP2 (RIA2020EF-2983-CSIGN: HPG). ERC Starting Grant (#757699: MQ). ERC (SG 757688: CJVA, KEA). This project has received funding from the European Union’s Horizon 2020 research and innovation programme - project EpiPose (101003688: AG, KLM, KP, WJE). This research was partly funded by the Global Challenges Research Fund (GCRF) project ‘RECAP’ managed through RCUK and ESRC (ES/P010873/1: CIJ). HDR UK (MR/S003975/1: RME). HPRU (This research was partly funded by the National Institute for Health Research (NIHR) using UK aid from the UK Government to support global health research. The views expressed in this publication are those of the author(s) and not necessarily those of the NIHR or the UK Department of Health and Social Care200908: NIB). MRC (MR/N013638/1: EF; MR/V027956/1: WW). Nakajima Foundation (AE). NIHR (16/136/46: BJQ; 16/137/109: BJQ, FYS; 1R01AI141534-01A1: DH; NIHR200908: AJK, LACC, RME; NIHR200929: CVM, NGD; PR-OD-1017-20002: AR, WJE). Royal Society (Dorothy Hodgkin Fellowship: RL). Singapore Ministry of Health (RP). UK DHSC/UK Aid/NIHR (PR-OD-1017-20001: HPG). UK MRC (MC_PC_19065 - Covid 19: Understanding the dynamics and drivers of the COVID-19 epidemic using real-time outbreak analytics: NGD, RME, SC, WJE; MR/P014658/1: GMK). UKRI (MR/V028456/1: YJ). Wellcome Trust (206250/Z/17/Z: AJK, TWR; 206471/Z/17/Z: OJB; 208812/Z/17/Z: SC; 210758/Z/18/Z: JDM, JH, SA, SFunk, SRM; 221303/Z/20/Z: MK). No funding (DCT, SH).

