## Supplementary material for "Optimising health and economic impacts of COVID-19 vaccine prioritisation strategies in the WHO European Region": Technical Appendix

**CMMID COVID-19 Working Group Members:**
Akira Endo, Gwenan M Knight, Joel Hellewell, Matthew Quaife, Oliver Brady, Rachael Pung, Yalda Jafari, Sam Abbott, Adam J Kucharski, Sebastian Funk, Rosalind M Eggo, W John Edmunds, Amy Gimma, Billy J Quilty, Samuel Clifford, James D Munday, Nikos I Bosse, Hamish P Gibbs, Nicholas G. Davies, Timothy W Russell, Christopher I Jarvis, Alicia Rosello, Kiesha Prem, Graham Medley, Simon R Procter, C Julian Villabona-Arenas, Damien C Tully, Katherine E. Atkins, Sophie R Meakin, Rachel Lowe, Kaja Abbas, Kathleen O'Reilly, Mihaly Koltai, William Waites, David Hodgson, Emilie Finch, Ciara V McCarthy, Paul Mee, Lloyd A C Chapman, Fiona Yueqian Sun, Stéphane Hué, Kerry LM Wong

**Figure S12. Optimal vaccine prioritisation strategies under different rollout scenarios and decision-making metrics using a longer vaccine waning period** ... 32

**7. Results of ordinal logistic regression exercise** ... **33**

**Figure S13. Coefficients and their corresponding 90% and 95% confidence interval in the ordinal logistic regression model.** ... 33

**8. [Sensitivity analysis] Underreporting** ... **34**

**Figure S14. Optimal vaccine prioritisation strategies under different roll-out scenarios and decision-making metrics considering underreporting** ... 34

**9. [Sensitivity analysis] Different decision time frames** ... **35**

**Figure S15. Optimal vaccine prioritisation strategies under different roll-out scenarios when decision-making metrics were summarised over different decision-making time frames.** ... 35

**10. Vaccinating adolescents** ... **36**

**Table S8. Counts of cases where a policy is ranked first in terms of optimising health and economic benefits (including ties)** ... 36

**11. Country-specific vaccine-prioritisation strategies by different vaccine profiles** ... **37**

**Figure S16. Optimal vaccine prioritisation strategies for different vaccine characteristics under R1 and R4.** ... 38

### 1. Details on Model Input & Assumptions

<table>
<tr><td colspan="5">Known Parameters based on Existing Knowledge</td></tr>
<tr><td>Index</td><td colspan="2">Variable</td><td>Values</td><td>Source</td></tr>
<tr><td>1</td><td colspan="2">Age-specific susceptibility</td><td>0.38 - 0.88</td><td>Davies et al.[1]</td></tr>
<tr><td>2</td><td colspan="2">Age-specific clinical progression rates</td><td>0.21 - 0.70</td><td>Davies et al.[1]</td></tr>
<tr><td>3</td><td colspan="2">Age-specific infection fatality rates</td><td>5e-6 - 0.13</td><td>Levin et al.[2]</td></tr>
<tr><td>4</td><td colspan="2">Age- and country-specific within-population contact pattern</td><td>Country-specific</td><td>Prem et al.[1]</td></tr>
<tr><td>5</td><td colspan="2">Country-specific population age structures</td><td>Country-specific</td><td>United Nations[3], see also 1.1</td></tr>
<tr><td>5</td><td colspan="2">Relationship between mobility and population contact pattern</td><td>Defined by linear and nonlinear functions for the workplace and other settings, respectively.</td><td>Davies et al.[4] by fitting to UK data.</td></tr>
<tr><td rowspan="5">6</td><td rowspan="5">CovidM</td><td>Latent period</td><td>~gramma (μ = 2.5, k = 2.5)</td><td rowspan="3">Pearson et al.[5]<br>Davies et al.[4]<br>Davies et al.[6]<br>Bi et al.[7]<br>Liu et al.[8]<br>Linton et al.[9]<br>Nishiura et al.[10]</td></tr>
<tr><td>Duration of preclinical infectiousness</td><td>~gramma (μ = 1.5, k = 4)</td></tr>
<tr><td>Duration of clinical infectiousness</td><td>~gramma (μ = 3.5, k = 4)</td></tr>
<tr><td>Duration of subclinical infectiousness</td><td>~gramma (μ = 5, k = 4)</td><td>Assumed</td></tr>
<tr><td>Mean duration of immunity from infection</td><td>3 years</td><td>Hall et al.[11]</td></tr>
</table>

**Table S1. Inputs and assumptions**

**Caption:** The variable index numbers correspond to their numberings in Figure 1 of the main text.

| Fitting Stage - input | | |
|---|---|---|
| Index | Variable | Source |
| 1 | Country-level daily COVID-19 Mortality (including 7-day rolling average) | Roser et al.[12]<br>Human Rights Watch[13] on change in mortality case definition in Kyrgyzstan/ Kazakhstan |
| 2 | Observed country-specific community mobility | Google LLC.[14] |
| 3 | COVID-19 Government Response Stringency Index and Government Response Tracker by country | Hale et al.[15] |

**Table S1. Inputs and assumptions (Continued)**

| Projection Stage - input and assumptions | | | | |
|---|---|---|---|---|
| Index | Variable | | Values | Source |
| 2 | Vaccine characteristics | Vaccine protection duration | Baseline = 52 weeks<br>Sensitivity analysis:<br>3 years | Assumed |
| | | Infection blocking efficacy | Baseline = 0.95, varied between 0 and 0.95 | |
| | | Disease blocking efficacy | Baseline = 0.95, varied between 0.5 and 0.95 | |
| | | Vaccine roll-out scenarios | 0.03 by mid-2021 and 0.2 by end of 2021, relatively slow roll-out may start after March 2021 | Gavi, the vaccine alliance[16], World Health Organisation[17,18] |
| | | Maximum willingness to receive vaccination | 0.7 for those between 20-59 and 0.9 for those above 90 | Wouter et al.[19]<br>Robinson et al.[20]<br>UK Government[21] |
| 3 | Impact and health economic metrics | Country-specific comorbidity Adjusted Life Expectancy | Age-specific | See 1.2 |
| | | Mean QALY associated with COVID-19 morbidity | 0.0307 | See 1.3 |
| | | Median QALD associated with AEFI | 1 | Oliver et al.[22] |
| | | Probability of adverse events following immunisation | 50% | Pfizer and BioNTech[23] by summing roughly summing |
| | | Country-specific GDP per capita | Country specific | See 1.4 |
| 4 | Projected mobility changes | | | GAM Model estimated<br><br>Association between mobility and contact is |

| | | | |
|---|---|---|---|
| | | | based on Davies et al.[4] |

**Table S1. Inputs and assumptions (Continued)**

### 1.1 Age pyramid in the WHO European Region

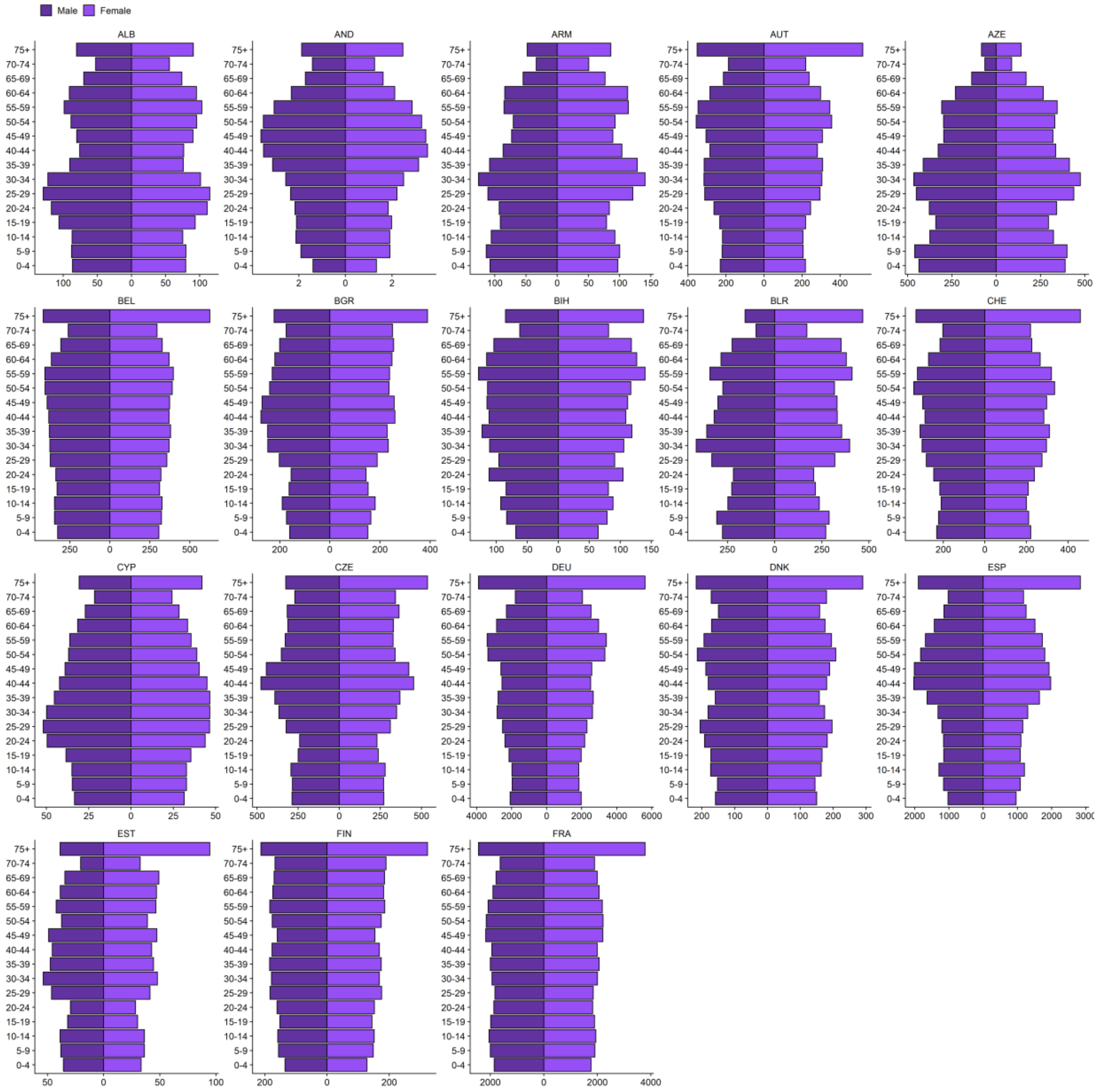

**Figure S1. Population age pyramid by country.**

Data source: United Nations.[3]

Caption: Countries are labelled using their three-digit World Bank country codes.

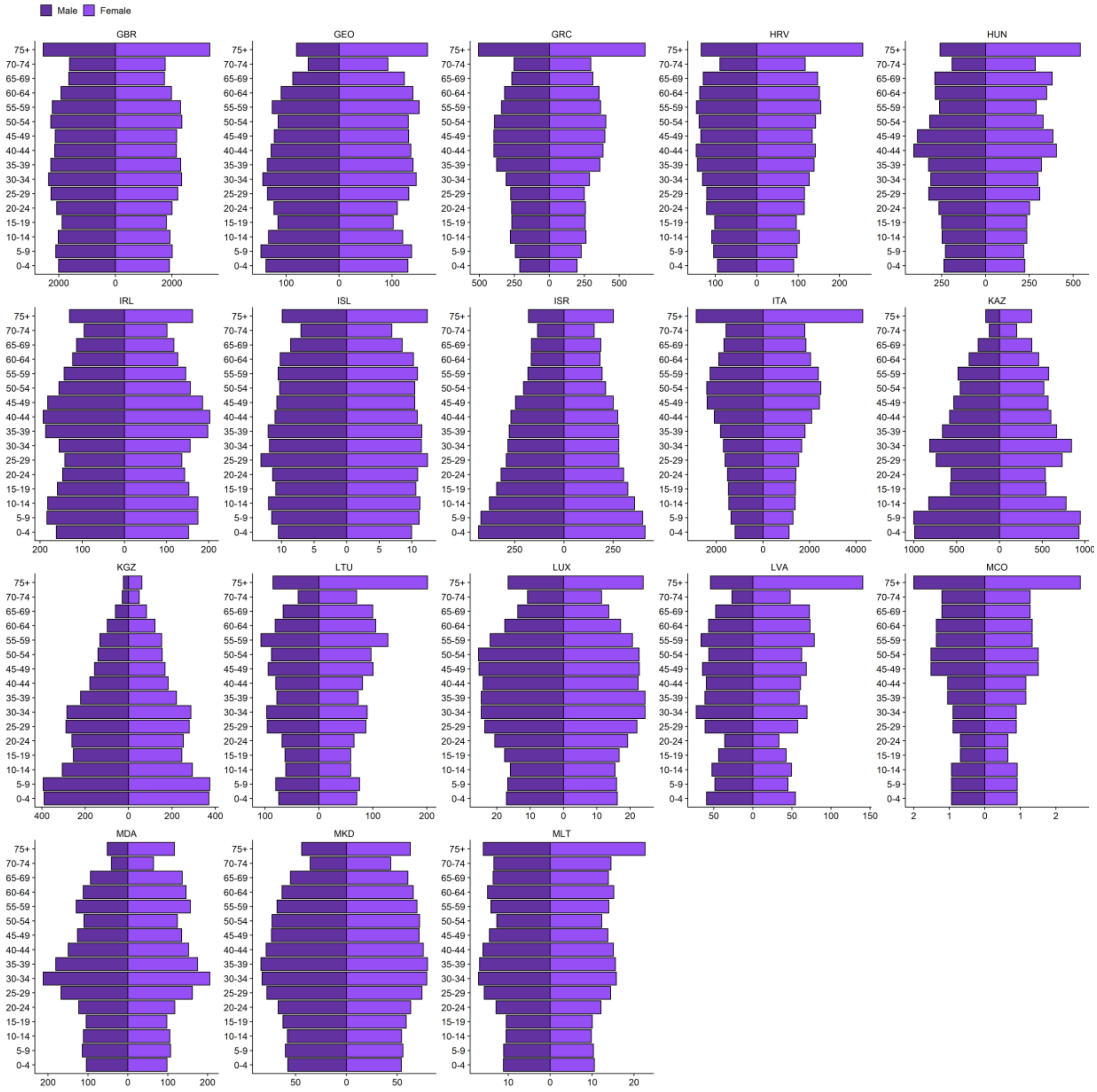

**Figure S1. Population age pyramid by country (continued).**

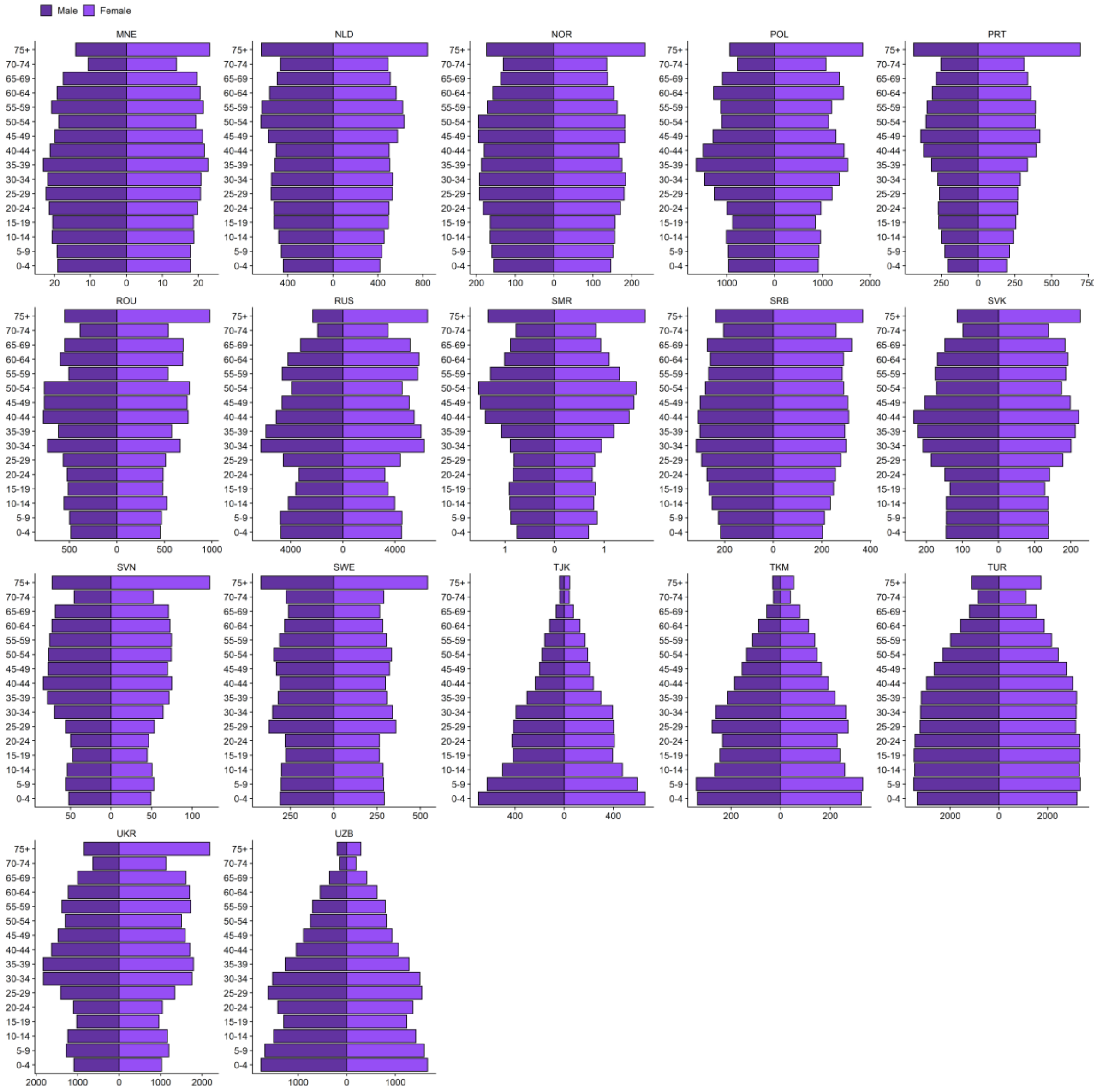

**Figure S1. Population age pyramid by country (continued).**

| Country Name | World Bank Country Code | Country Name | World Bank Country Code |
|---|---|---|---|
| Albania | ALB | Lithuania | LTU |
| Andorra | AND | Luxembourg | LUX |
| Armenia | ARM | Malta | MLT |
| Austria | AUT | Monaco | MCO |
| Azerbaijan | AZE | Montenegro | MNE |
| Belarus | BLR | Netherlands | NLD |
| Belgium | BEL | Norway | NOR |
| Bosnia & Herzegovina | BIH | Poland | POL |
| Bulgaria | BGR | Portugal | PRT |
| Croatia | HRV | Moldova | MDA |
| Cyprus | CYP | Romania | ROU |
| Czechia | CZE | Russia | RUS |
| Denmark | DNK | San Marino | SMR |
| Estonia | EST | Serbia | SRB |
| Finland | FIN | Slovakia | SVK |
| France | FRA | Slovenia | SVN |
| Georgia | GEO | Spain | ESP |

| | | | |
|---|---|---|---|
| Germany | DEU | Sweden | SWE |
| Greece | GRC | Switzerland | CHE |
| Hungary | HUN | Tajikistan | TJK |
| Iceland | ISL | North Macedonia | MKD |
| Ireland | IRL | Turkey | TUR |
| Israel | ISR | Turkmenistan | TKM |
| Italy | ITA | Ukraine | UKR |
| Kazakhstan | KAZ | United Kingdom | GBR |
| Kyrgyzstan | KGZ | Uzbekistan | UZB |
| Latvia | LVA | | |

**Table S2. Country names and their corresponding World Bank country code.**

#### 1.2. Comorbidity-adjusted life expectancy, comorbidity- and quality-adjusted life expectancy, and discounted life expectancy

Data on life expectancies for each country in the WHO European Region were taken from the website of WHO.[24] We adjusted the life expectancies for higher risks of death due to comorbidities in those who die from COVID-19 using a recently proposed method and assuming an increased risk of 50%.[25] We also adjusted for health-related quality of life (HRQoL) by age using EQ-5D-3L population norms from the seven countries in Europe with a time-trade off value set available (i.e., Denmark, France, Germany, Italy, Netherlands, Spain, UK),[26] which indicates the potential loss of health-related quality of life (HRQoL) due to death per country (as deaths are based on country-specific estimates). Lastly, we also accounted for time preferences using a discount rate of 3.0% for future years.[27]

#### 1.3. QALY associated with COVID-19 morbidity

For morbidity, we assumed for each non-hospitalised case a QALY loss equivalent to symptomatic episodes of pandemic influenza-like illness.[28] Furthermore, we assumed that 10% of cases are hospitalised (based on raw data of hospitalisations to cases), losing 0.0201 QALYs for more than 2 months post-discharge.[29] Of the hospitalised cases, 50% were assumed to survive treatment in intensive-care units,[30] with an estimated longer-term impact of ICU survivors of 0.15 QALYs.[31,32] Another 10% of non-hospitalised cases were assumed to suffer from post-acute symptoms (long COVID),[33] for whom we assumed a similar impact comparable to ICU survivors of 0.15 QALYs lost. In total, each symptomatic case is thus assigned a health loss equivalent to 0.0307 QALYs.

#### 1.4. GDP per capita used in the human capital approach

For the human capital approach, we used the annual GDP per capita in international dollars (intl$) in 2019 (or 2018 if unavailable) from the World Bank, converted by purchasing power parity (PPP).[34] In the absence of these data for Andorra and Monaco, we used their GDP per capita in current US$ (without PPP conversion) from 2019 and 2018, respectively. With the GDP being a country-level productivity measure, combined with the country-specific life expectancies we derive an estimate of the economic losses per country (Figure S4) to add dimension to the health impact.

### 2. Projecting human contacts

#### 2.1. Linking stringency indices to community mobility

We projected human mobility after March 2021 and before December 2022 (during which observation is not yet available) using a general additive model:

Mobility ~ day of week + (1 | country) + setting * day of week + mobility setting * month of year + stringency index

In which:

| Symbol | Values | Notes |
|---|---|---|
| day of week | Nominal categorical values 1-7 | To capture within week variability. Work-related variability, for example, vary tremendously depending on the day of week |
| 1 \| country | Country code | To capture country-specific random effects |
| setting | Type of mobility | Type of mobility, one of “retail”, “transit”, “grocery”, and “work”. |
| setting * day of week | The interaction term between the type of mobility and day of week | To capture the interaction between type of mobility and day of the week. |
| setting * month of year | The interaction term between the type of mobility and month of year | To capture variable specific mobility seasonality. This has similar problems with using day of year as a predictor - 2020 has not fully elapsed yet so we aren’t sure what happens in Jan or Dec. In this study, we assume Dec to be similar to Nov, and Jan to be similar to Feb. |
| stringency index^ | Government stringency index describing the intensity of COVID-19 related non-pharmaceutical interventions | To capture the large decrease in early 2020. Without adjusting for the stringency index, the early year mobility for 2021 and 2020 may be artificially pulled lower. |

**Table S3. Covariates table for the general additive model (GAM) used in mobility projection (projection stage)**

**Caption:** Based on COVID-19 related non-pharmaceutical interventions recorded in Oxford COVID-19 Government Response Tracker, the stringency index represented the extent of containment and closure policies and public health information campaigns on the country level.[15] The value of the stringency index ranges between 0 and 100, with 0 indicating the least stringent conditions, and 100 the most stringent conditions. We extracted stringency index data on 5 Mar 2021, on which complete records for our countries of interest are available before 22 Feb 2021.

#### 2.2. Projecting stringency indices

| Countries | Min | 1st Quarter | Median | 3rd Quarter | Max |
|---|---|---|---|---|---|
| All available (n = 50) | 22 | 54 | 61 | 71 | 88 |
| Countries with fitted models (n = 38) | 28 | 56 | 68 | 72 | 88 |

**Table S4. Stringency indices on at the end of the observation window (i.e., 22 Feb 2021)**

We assume over one year from 22 Feb 2021, as vaccines roll out, stringency indices gradually recover towards pre-pandemic levels. However, due to long-term behaviour and policy changes, we expect that stringency indices will never fully return to 0 in the time frame of this study. Thus, we assume stringency indices will return to 10 over 365 days, regardless of their positions on 22 Feb 2021, following a sigmoid function. After 22 Feb 2022, the stringency indices would stay at 10 to reflect any long-term changes COVID-19 policies have on human behaviours. We impute the mobility for countries without mobility data by taking an average of the geographic neighbours.

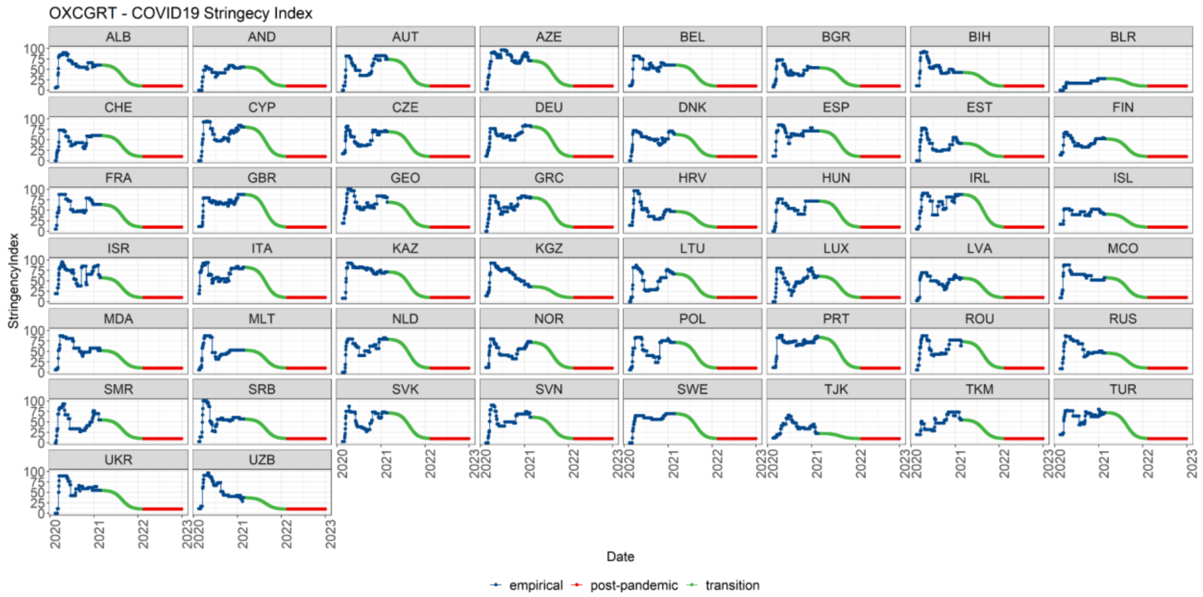

**Figure S2. Stringency indices by country after incorporating the assumption on mobility recovery.**

**Caption:** Dark blue – empirical observations; green – transition phase; red – post-pandemic re-stabilised phase.

| Countries | Number of Countries with Community Mobility Report | Number of Countries with Stringency Indices |
|---|---|---|
| All (n = 53) | 42 | 50 |
| Countries with fitted models (n = 38) | 35 | 36 |

**Table S5. Missingness in community mobility reports and stringency indices.**

Of all countries we fitted, Albania (ALB), Armenia (ARM), and Azerbaijan (AZE) do not appear in community mobility reports; Armenia (ARM) and North Macedonia (MKD) cannot be found in the stringency index database.

#### 2.3. Projected population contacts

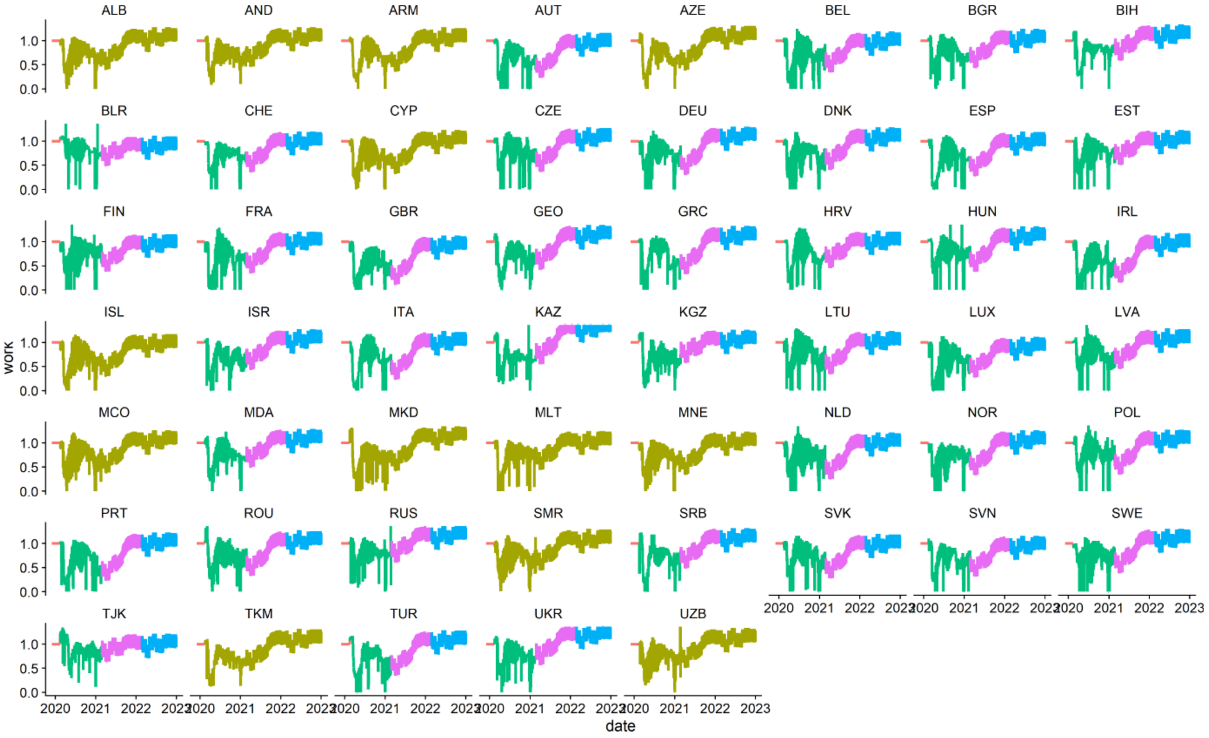

**Figure S3. Projected multipliers of daily contacts before December 2022 in the *work* setting**

**Caption:** Projected multipliers of baseline contact matrices capture the changes in population contact intensity. These multipliers were calculated using projected stringency indices, projected community mobility, and the relationship between daily contacts and community mobility defined by Davies et al.[4] using UK data. Colors represent: coral = pre-pandemic baseline; dark yellow = imputed data for countries where community mobility data is not available; green = empirical observation; pink = transition phase between pandemic and post-pandemic phases; blue = post-pandemic phase.

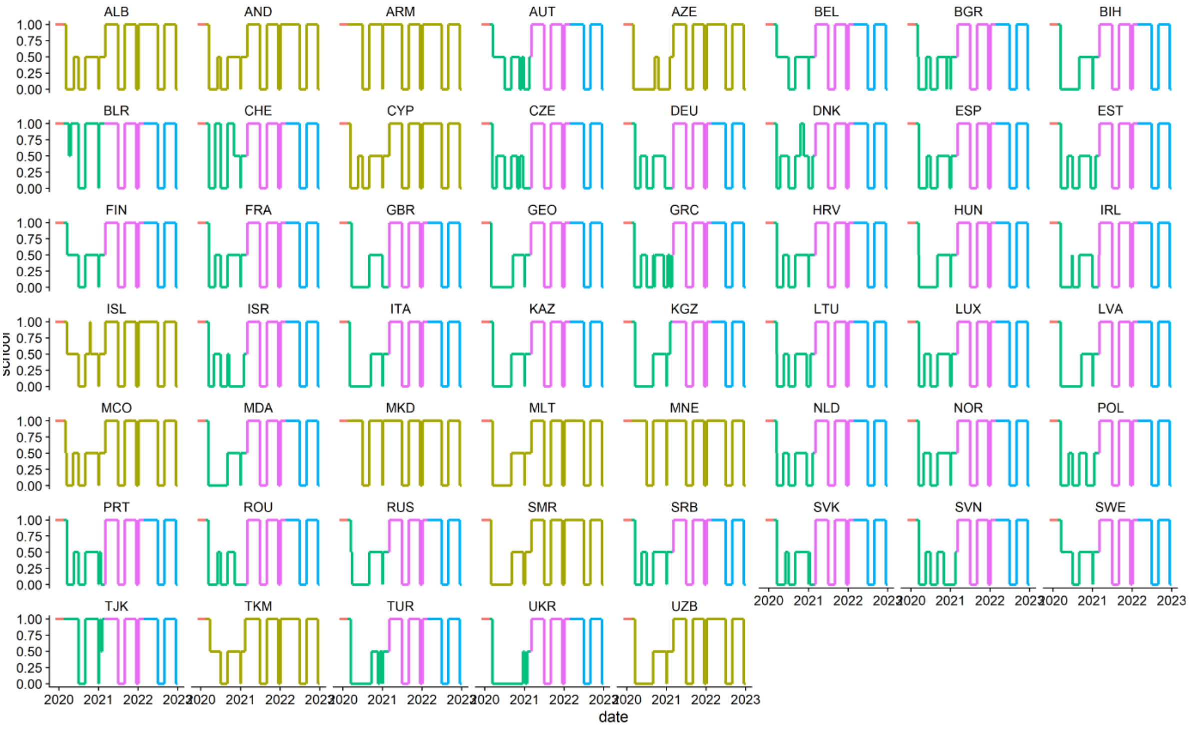

**Figure S4. Projected multipliers of daily contacts before December 2022 in the *school* setting.**

**Caption:** Projected multipliers of baseline contact matrices captured the changes in population contact intensity. Colors represent: coral = pre-pandemic baseline; dark yellow = imputed data for countries where community mobility data is not available; green = empirical observation; pink = transition phase between pandemic and post-pandemic phases; blue = post-pandemic phase. During the period where empirical observation was available, the contacts in the *school* setting were modulated by the “school closure” variable (i.e. C1_school_closing) in the Oxford COVID-19 government response tracker. The variable C1_school_closing is ordinal: when C1_school_closing reached the highest value (i.e. 3, require closing all levels), the multiplier was set to 0 to indicate no school contacts; when C1_school_closing reached the lowest value (i.e. 0, require closing all levels), the multiplier was set to 1 to indicate *school*-based contacts were similar to pre-pandemic levels; when C1_school_closing was set to levels in between, the multiplier was assumed to be 0.5 to indicate an intermediate level of *school*-based contact intensity. During the transition between pandemic and post-pandemic phases and the post-pandemic phases, contacts in the *school* setting were only driven by the summer and winter holidays. The timing of summer and winter holidays varies in the European Union.[35] Here, we broadly assume summer holidays to go for two months between July and August, and winter holidays to go for 3 weeks between mid-December to the first week of January.  During school holidays, *school*-based contacts were set to 0.

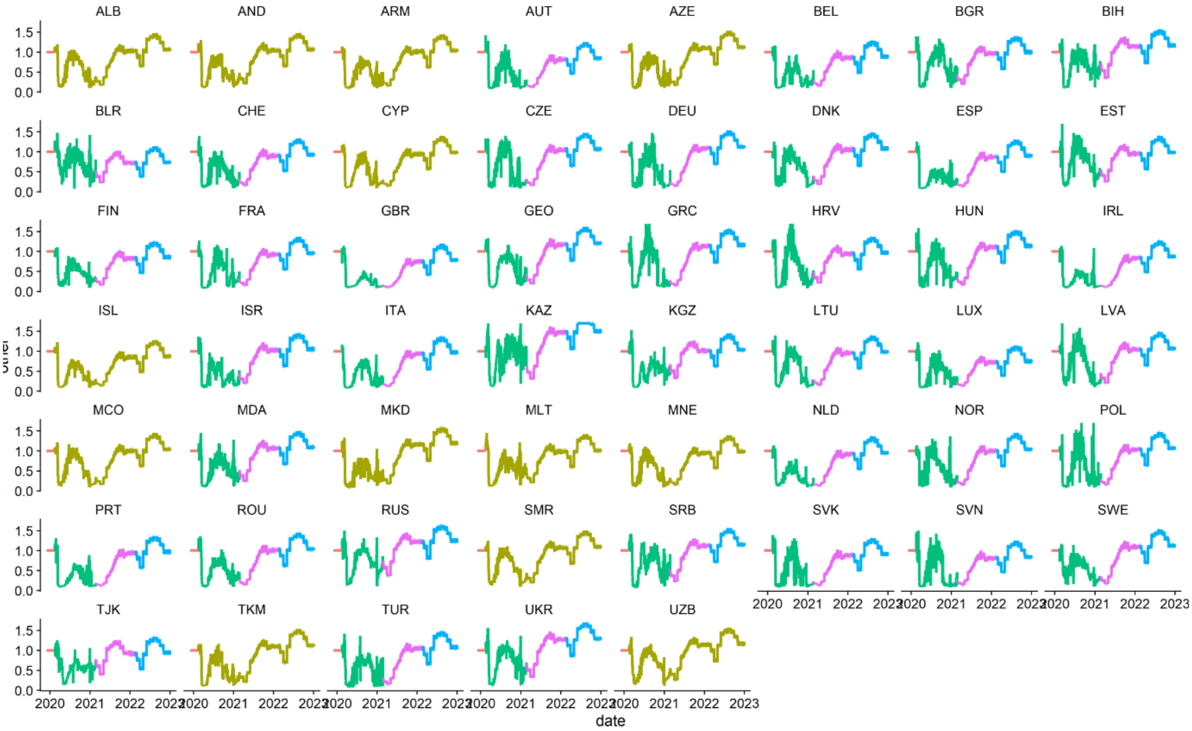

**Figure S5. Projected multipliers of daily contacts before December 2022 in the *others* setting.**

**Caption:** Projected multipliers of baseline contact matrices capture the changes in population contact intensity. These multipliers were calculated using projected stringency indices, projected community mobility, and the relationship between daily contacts and community mobility defined by Davies et al.[4] using UK data. Colors represent: coral = pre-pandemic baseline; dark yellow = imputed data for countries where community mobility data is not available; green = empirical observation; pink = transition phase between pandemic and post-pandemic phases; blue = post-pandemic phase.

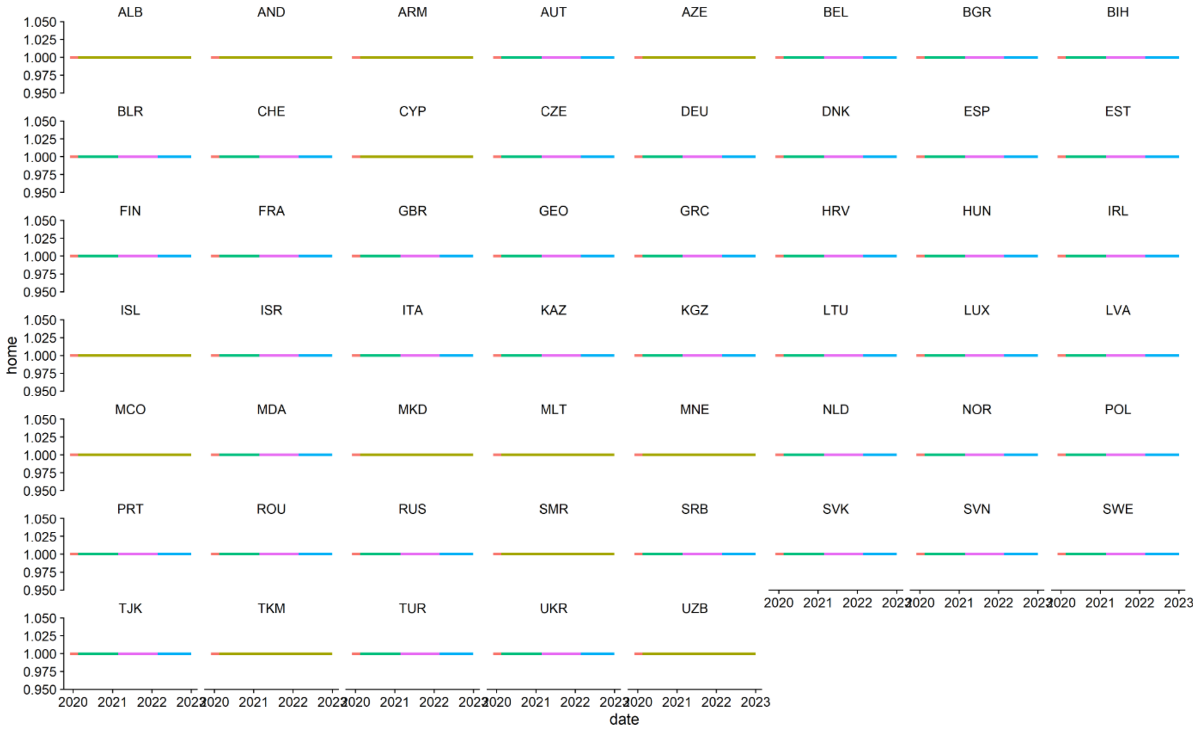

**Figure S6. Projected multipliers of daily contacts before December 2022 in the *home* setting.**

**Caption:** Projected multipliers of baseline contact matrices capture the changes in population contact intensity. Colors represent: coral = pre-pandemic baseline; dark yellow = imputed data for countries where community mobility data is not available; green = empirical observation; pink = transition phase between pandemic and post-pandemic phases; blue = post-pandemic phase. Home-based contacts are expected to stay constant in this study. The Google community mobility report showed increased time spent at home during the pandemic.[14] We argue that this change does not affect transmission as both pandemic and pre-pandemic time and contacts at home are likely above the time and contacts required for transmission.[36]

### 3. Impact and Health Economic Metrics

The three health-economic metrics can then be calculated using the following equations:

(1) Comorbidity-adjusted life expectancy (cLE) loss =
age-specific COVID-19 mortality *
age-specific comorbidity-adjusted life expectancy (adjLE)

(2) Comorbidity- and quality-adjusted life year (cQALY) loss =
age-specific discounted comorbidity- and quality- adjusted life expectancy (adjQALEdisc) *
age-specific COVID-19 mortality +
mean QALY loss associated with COVID-19 morbidity *
COVID-19 symptomatic cases +
mean QALY loss associated with AEFI * Number of vaccines deployed *
AEFI occurrence probability

(3) Human Capital (HC) loss =
total discounted life expectancy (LEdisc) * GDP per capita

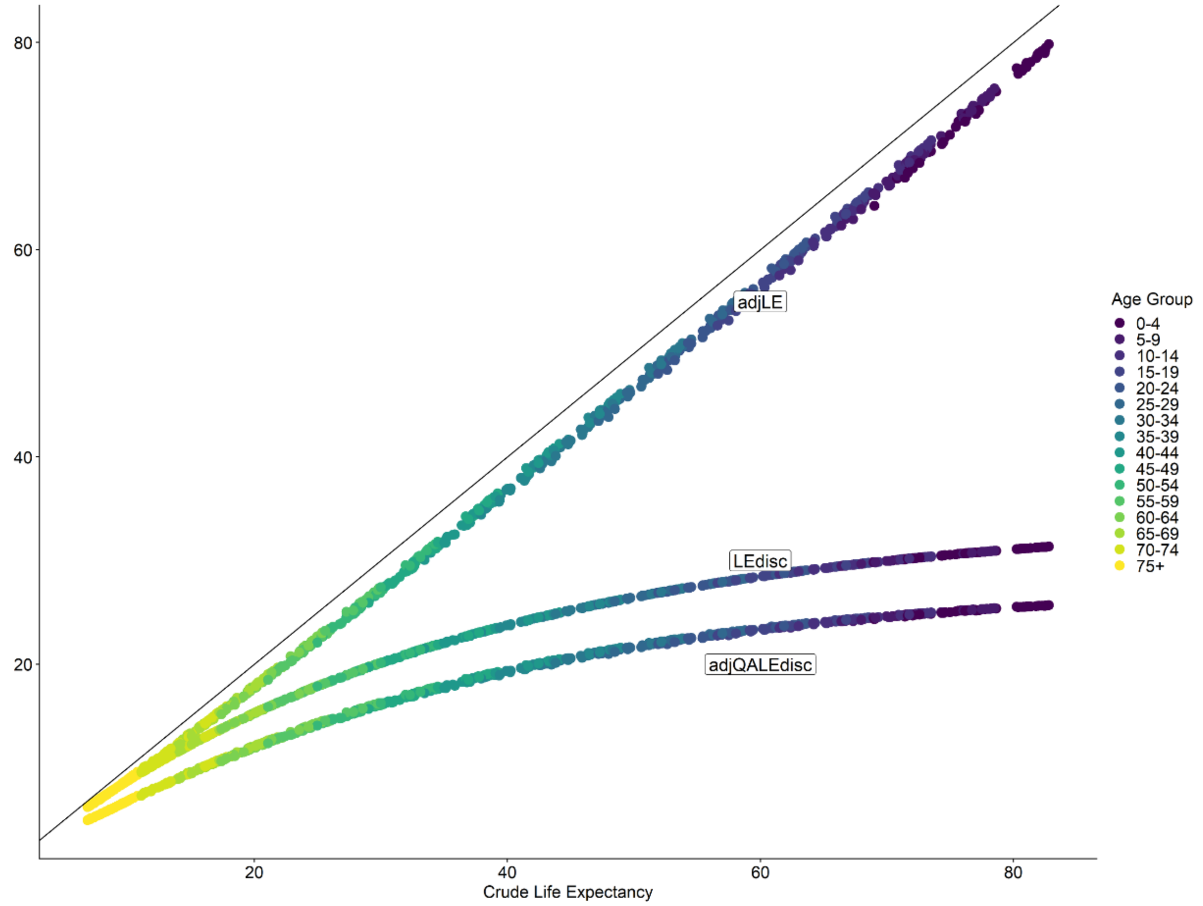

**Figure S7. Numeric differences between crude life expectancy and comorbidity adjusted life expectancy (adjLE), discounted life expectancy (LEdisc) and discounted comorbidity- and quality-adjusted life expectancy (adjQALEdisc).**

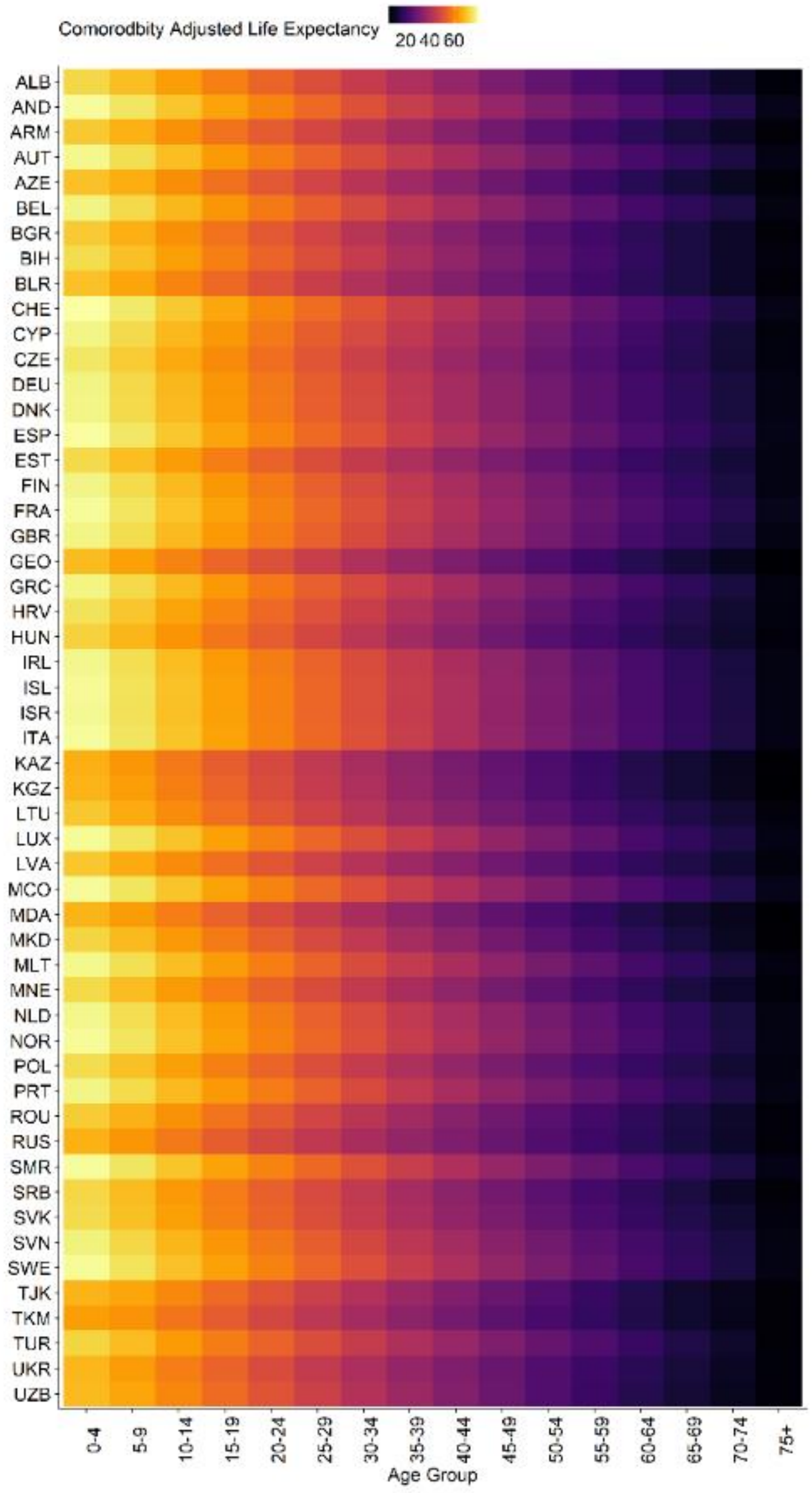

**Figure S8. Comorbidity adjusted life expectancy by age and by country.**

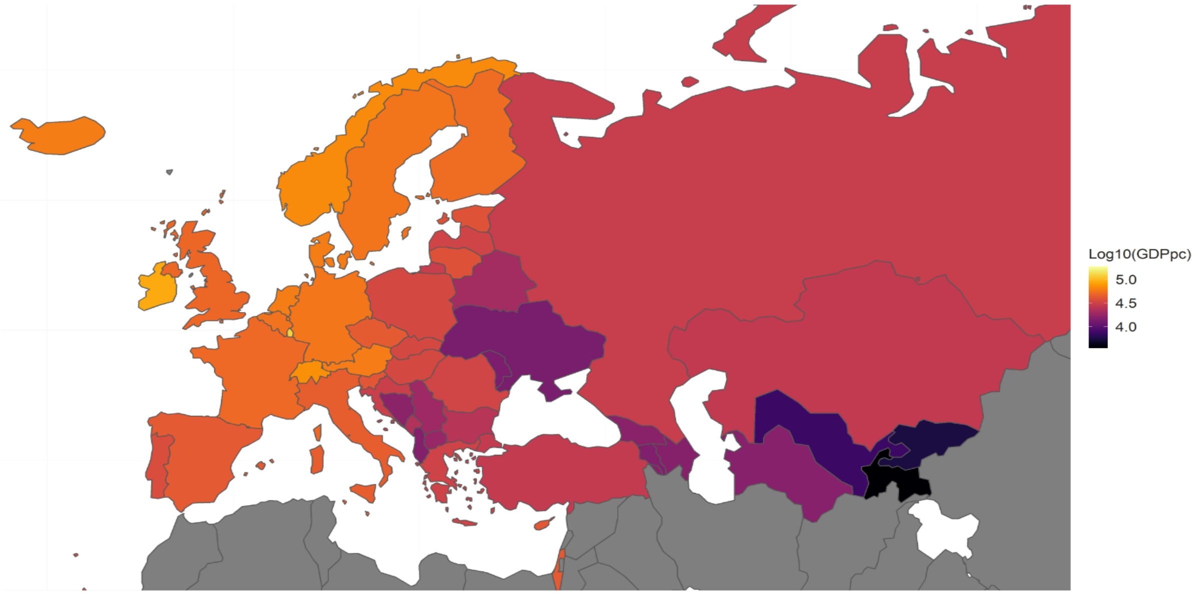

**Figure S9. Gross Domestic Production per capita (GDPpc) in the WHO European Region**

### 4. Additional results from the fitting process

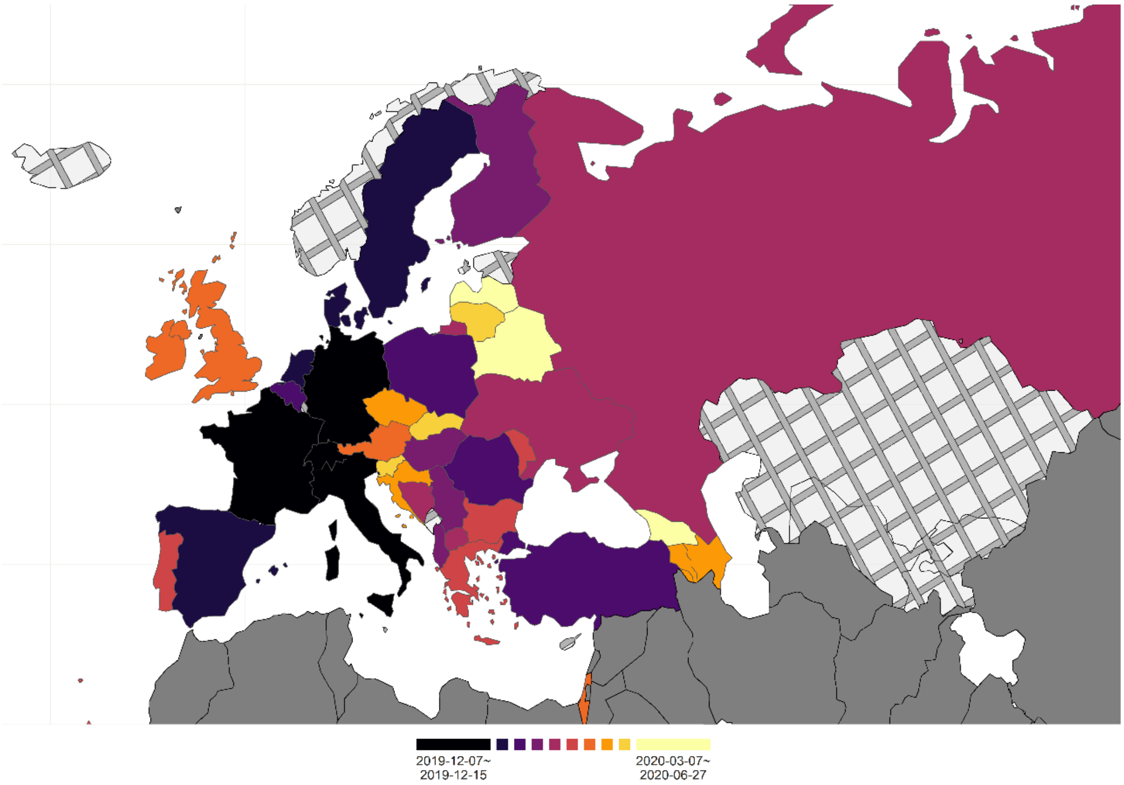

**Figure S10. Fitted infection introduction dates in the WHO European Region.**

**Caption**: The underlying fitting structure involves two varying parameters - infection introduction date and the basic reproduction number. Country shapefiles are downloaded from Eurostat GISCO.[37] Cross-hatched regions indicate countries where model fit was not achieved due to data availability and quality issues discussed in the main text.

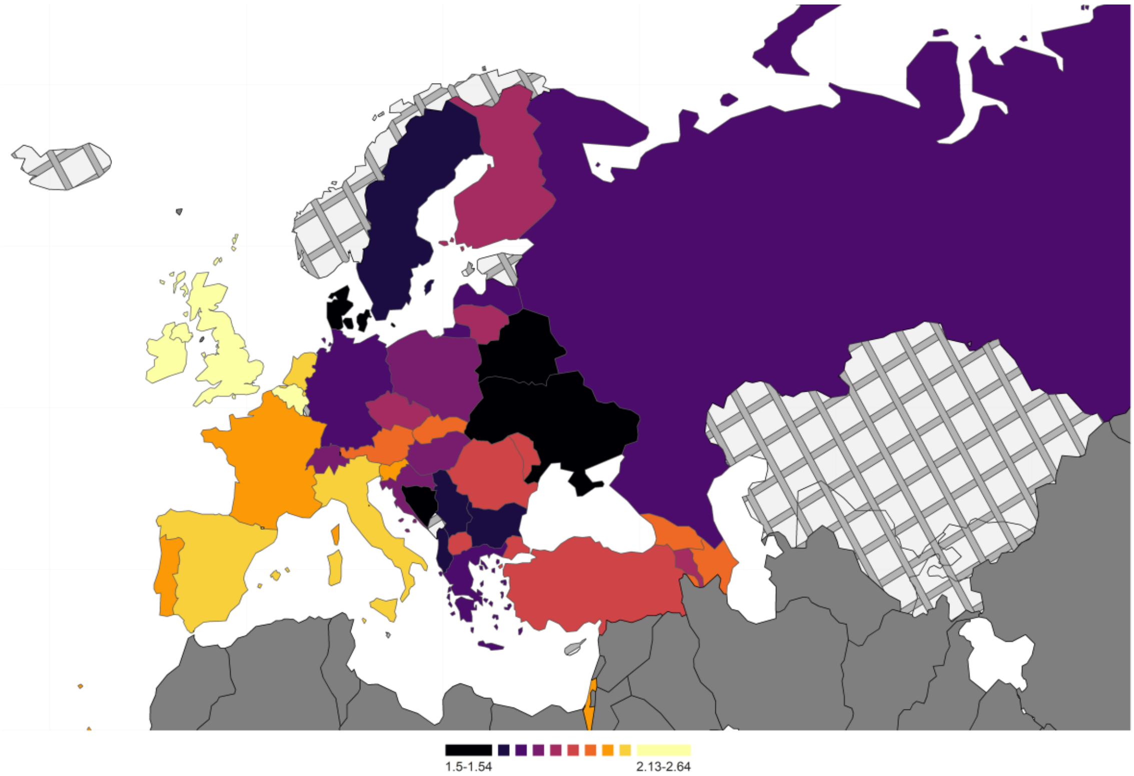

**Figure S11. Fitted basic reproduction numbers in the WHO European Region.**

Caption: The underlying fitting structure involves two varying parameters - infection introduction date and the basic reproduction number. Country shapefiles are downloaded from Eurostat GISCO.[37] Cross-hatched regions indicate countries where model fit was not achieved due to data availability and quality issues discussed in the main text.

### 5. Proportions of countries and populations by optimal vaccine prioritization strategies.

| Roll-out Scenario | Decision-making Metrics | Proportion of countries with fitted models with this optimal vaccine prioritisation strategy (n = 38) | | | |
|---|---|---|---|---|---|
| | | V+ | V20 | V60 | V75 |
| R1 | Deaths | 0.0263 | 0.1842 | 0.1316 | **0.6579** |
| R1 | Cases | 0.0000 | **1.0000** | 0.0000 | 0.0000 |
| R1 | Adj. Life Expectancy | 0.0263 | 0.3684 | **0.5526** | 0.0526 |
| R1 | Quality Adj. Life Years | 0.0526 | 0.3947 | **0.5000** | 0.0526 |
| R1 | Human Capital | 0.0263 | 0.3421 | **0.6053** | 0.0263 |
| R2 | Deaths | 0.2105 | **0.3158** | 0.2368 | 0.2368 |
| R2 | Cases | 0.1053 | **0.8947** | 0.0000 | 0.0000 |
| R2 | Quality Adj. Life Years | 0.2895 | **0.5263** | 0.1842 | 0.0000 |
| R2 | Adj. Life Expectancy | 0.2632 | **0.5789** | 0.1579 | 0.0000 |
| R2 | Human Capital | 0.2632 | **0.4737** | 0.2632 | 0.0000 |
| R3 | Deaths | 0.3421 | 0.1579 | **0.4474** | 0.0526 |
| R3 | Cases | **0.4474** | 0.0526 | 0.2895 | 0.2105 |
| R3 | Adj. Life Expectancy | **0.3684** | 0.1579 | 0.3421 | 0.1316 |

| | | | | | |
|---|---|---|---|---|---|
| R3 | Quality Adj. Life Years | **0.3947** | 0.0789 | 0.4211 | 0.1053 |
| R3 | Human Capital | **0.3684** | 0.1579 | **0.3684** | 0.1053 |
| R4 | Deaths | 0.4474 | **0.5000** | 0.0263 | 0.0263 |
| R4 | Cases | **0.4737** | 0.1316 | 0.1316 | 0.2632 |
| R4 | Adj. Life Expectancy | **0.4737** | 0.4474 | 0.0789 | 0.0000 |
| R4 | Quality Adj. Life Years | **0.5000** | 0.4211 | 0.0789 | 0.0000 |
| R4 | Human Capital | **0.4737** | 0.4737 | 0.0526 | 0.0000 |

**Table S6. Proportions of countries with specific optimal vaccine prioritization strategies.**

**Caption:** The denominator for these proportions is 38, the number of countries within the WHO European Region without data availability or sparsity issues.

| Roll-out Scenario | Decision-making Metrics | Proportion of population in countries with fitted models with this optimal vaccine prioritisation strategy (n = 848,407) | | | |
|---|---|---|---|---|---|
| | | V+ | V20 | V60 | V75 |
| R1 | Deaths | 0.0126 | 0.1521 | 0.0499 | **0.7854** |
| R1 | Cases | 0 | **1** | 0 | 0 |
| R1 | Adj. Life Expectancy | 0.0126 | 0.2454 | **0.7362** | 0.0057 |
| R1 | Quality Adj. Life Years | 0.132 | 0.2581 | **0.6042** | 0.0057 |
| R1 | Human Capital | 0.0126 | 0.2331 | **0.752** | 0.0022 |
| R2 | Deaths | 0.1721 | 0.1169 | 0.316 | **0.395** |
| R2 | Cases | 0.134 | **0.866** | 0 | 0 |
| R2 | Quality Adj. Life Years | 0.2419 | **0.4352** | 0.323 | 0 |
| R2 | Adj. Life Expectancy | 0.2467 | **0.5017** | 0.2517 | 0 |
| R2 | Human Capital | 0.2405 | **0.3988** | 0.3607 | 0 |
| R3 | Deaths | **0.474** | 0.1328 | 0.2911 | 0.1021 |
| R3 | Cases | **0.5175** | 0.0213 | 0.2189 | 0.2423 |
| R3 | Adj. Life Expectancy | **0.4788** | 0.1328 | 0.2404 | 0.148 |

| | | | | | |
|---|---|---|---|---|---|
| R3 | Quality Adj. Life Years | **0.4948** | 0.032 | 0.3455 | 0.1278 |
| R3 | Human Capital | **0.4788** | 0.1328 | 0.2606 | 0.1278 |
| R4 | Deaths | 0.2541 | **0.7401** | 0.0022 | 0.0035 |
| R4 | Cases | 0.2576 | **0.3194** | 0.2465 | 0.1764 |
| R4 | Adj. Life Expectancy | 0.2606 | **0.6568** | 0.0827 | 0 |
| R4 | Quality Adj. Life Years | 0.2641 | **0.6465** | 0.0895 | 0 |
| R4 | Human Capital | 0.2606 | **0.7337** | 0.0057 | 0 |

**Table S7. Proportions of populations with specific optimal vaccine prioritization strategies.**

**Caption:** The denominator for these proportions is 848,407, the populations of countries within the WHO European Region without data availability or sparsity issues.

### 6. [Sensitivity analysis] Longer waning period for vaccine-induced immunity

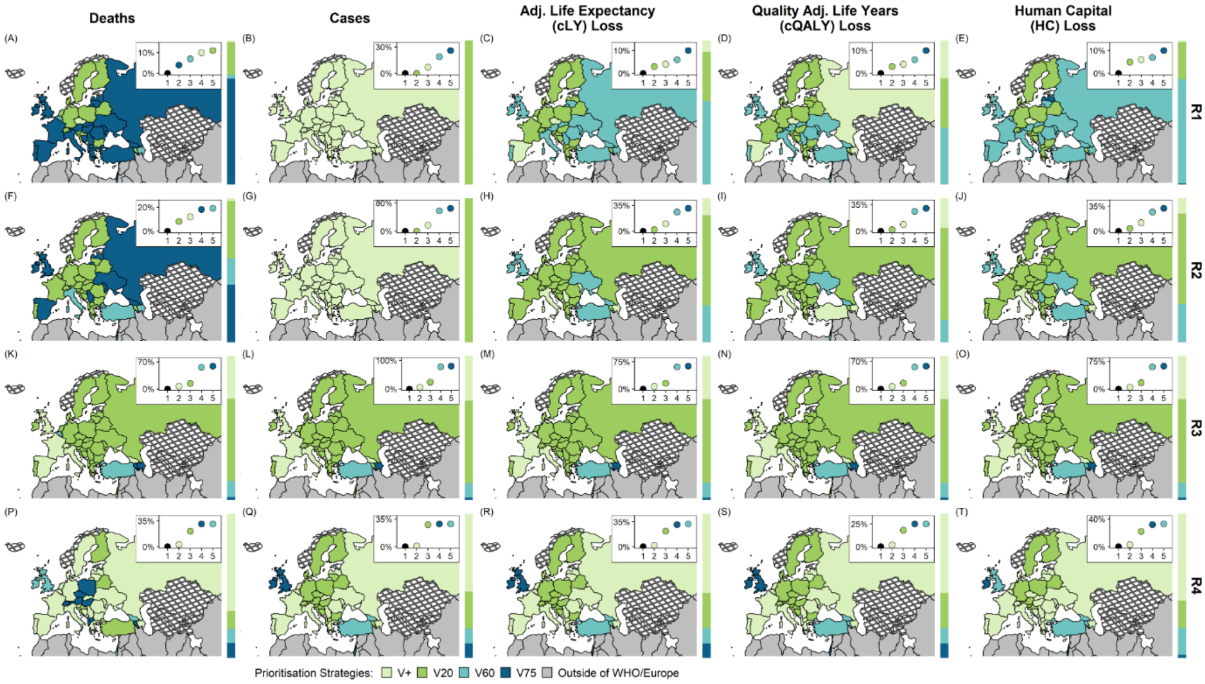

**Figure S12. Optimal vaccine prioritisation strategies under different rollout scenarios and decision-making metrics using a longer vaccine waning period**

**Caption:** The underlying fitting structure involves two varying parameters - infection introduction dates and the basic reproduction numbers. Vaccine-induced immunity is assumed to wane exponentially over 3 years (as opposed to 52 weeks, presented in the main text). Main panel - Optimal strategies across the WHO European Region identified using decision metrics of cumulative COVID-19 deaths, cases, and losses in adjusted life expectancy (cLE), quality-adjusted life-years (cQALY), and human capital (HC). Inner panel - Comparing the use of a given prioritisation strategy across the WHO European Region against the use of country-specific optimal prioritisation strategies (indicated with black points). Sidebars - The proportion of total population for which each prioritisation strategy is optimal. Country shapefiles are downloaded from Eurostat GISCO;[37] countries marked by crosshatch patterns are those that were not included in the fitting stage.

### 7. Results of ordinal logistic regression exercise

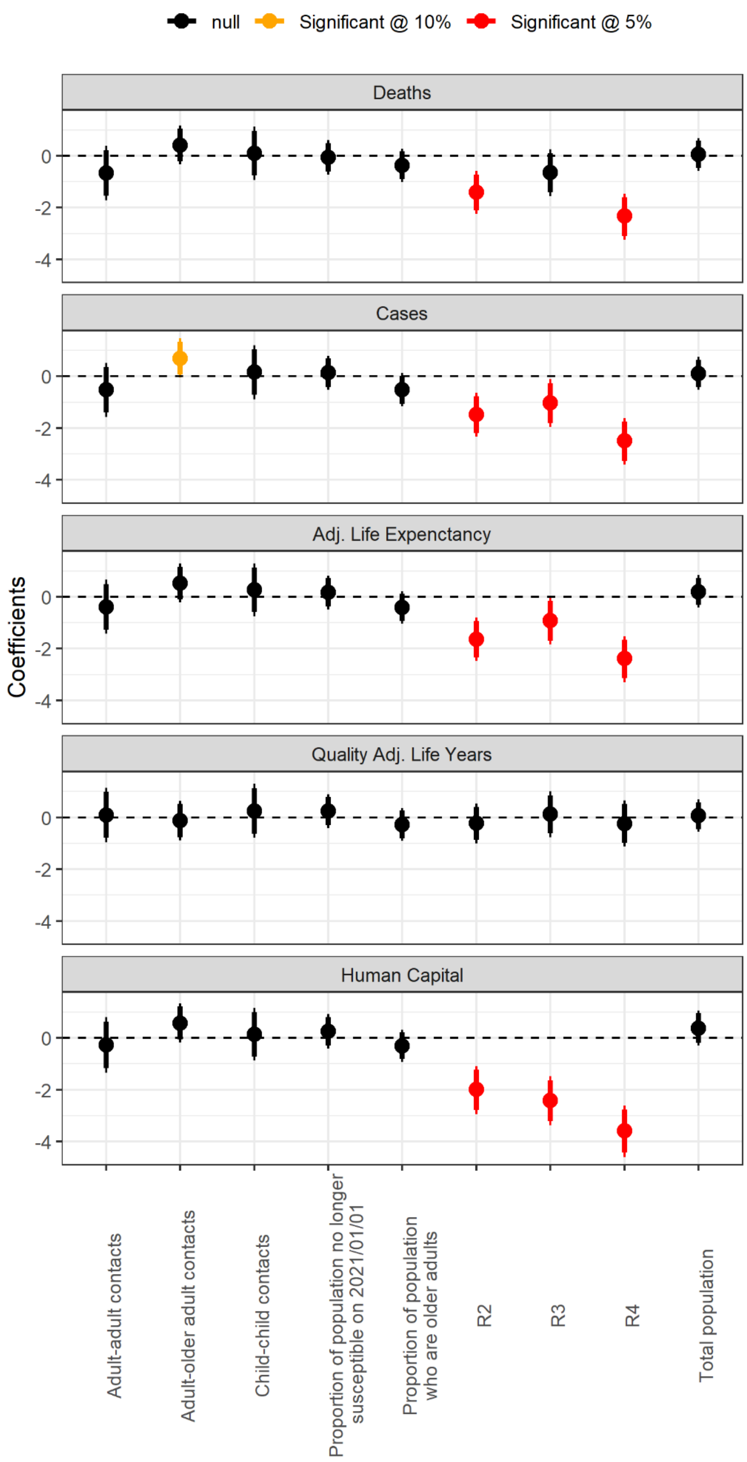

**Figure S13. Coefficients and their corresponding 90% and 95% confidence interval in the ordinal logistic regression model.**

**Caption**: The sole dependent variable is the optimal vaccine prioritisation strategy identified. There are five groups of independent variables: (1) age-specific contact patterns, (2) vaccine rollout scenarios, (3) population size, (4) proportion of older adults, and (5) the proportion of individuals no longer susceptible to SARS-CoV-2 by 01 January 2021. Not all variables within each group were presented here - those with Pearson's correlation larger than 0.4 were eliminated to avoid multicollinearity issues.

### 8. [Sensitivity analysis] Underreporting

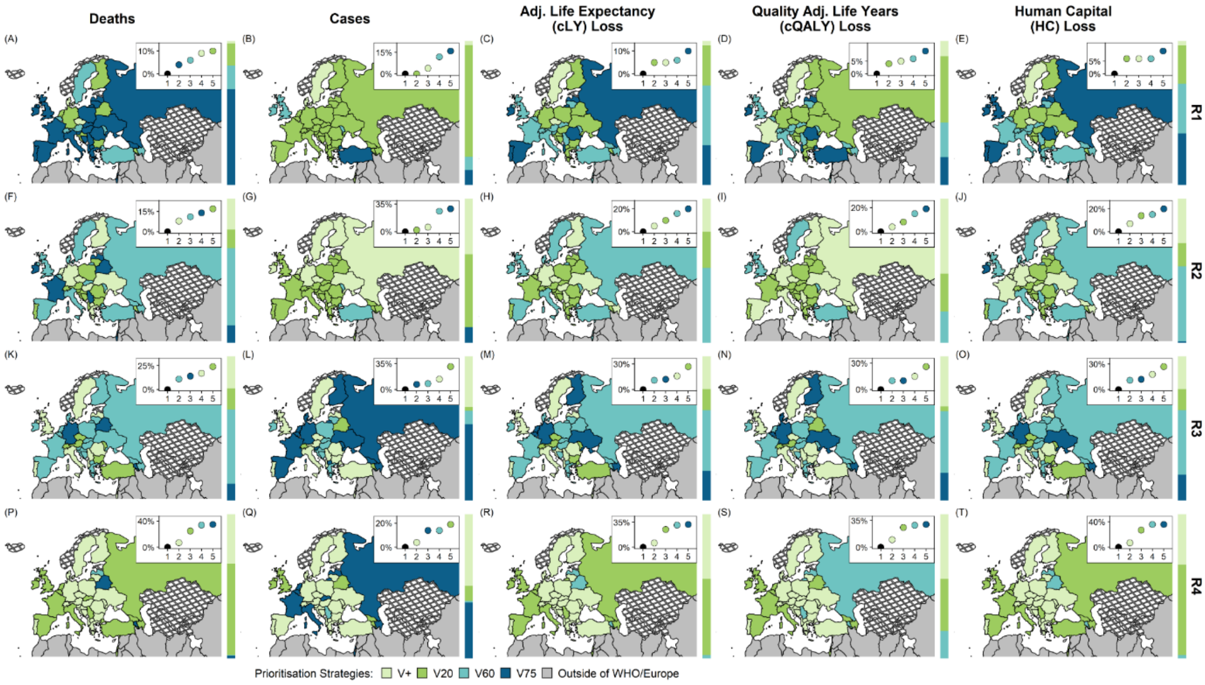

**Figure S14. Optimal vaccine prioritisation strategies under different roll-out scenarios and decision-making metrics considering underreporting**

**Caption:** The underlying fitting structure involves three varying parameters - infection introduction dates, the basic reproduction numbers, and an underreporting probability. Main panel - Optimal strategies across the WHO European Region identified using decision metrics of cumulative COVID-19 deaths, cases, and losses in adjusted life expectancy (cLE), quality-adjusted life-years (cQALY), and human capital (HC). Inner panel - Comparing the use of a given prioritisation strategy across the WHO European Region against the use of country-specific optimal prioritisation strategies (indicated with black points). Sidebars - The proportion of total population for which each prioritisation strategy is optimal. Country shapefiles are downloaded from Eurostat GISCO;[37] countries marked by crosshatch patterns are those that were not included in the fitting stage.

### 9. [Sensitivity analysis] Different decision time frames

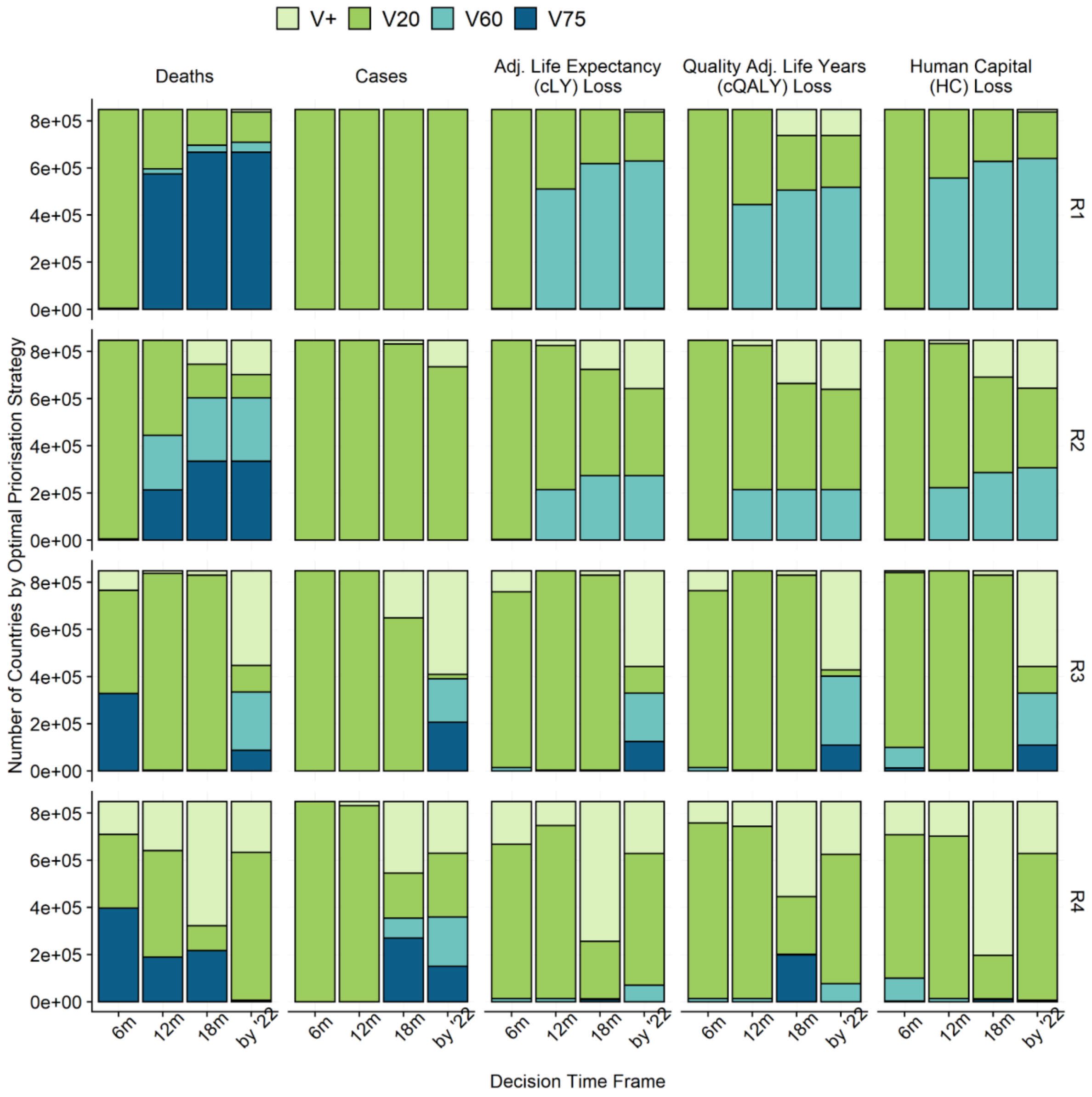

**Figure S15. Optimal vaccine prioritisation strategies under different roll-out scenarios when decision-making metrics were summarised over different decision-making time frames.**

**Caption:** The right columns in each panel is another way to visualise the results already presented in the main text, Figure 4 - the results presented in these columns are each decision-making metric summarised between 01 Jan 2021 and 31 Dec 2022. Noticeably, the vaccination programs elapsed for different durations under different rollout scenarios. For R1 and R2, the vaccination campaigns were assumed to start on 01 March 2021; for R3 and R4, the vaccination campaigns were assumed to start on 01 January 2021. The rest represents results summarised over decision time frames. Among them, "6m", "12m" and "18m" represents decision time frames ending on the 6th, 12th, and 18th months after the start of vaccination campaigns. In these columns, vaccine program lengths were the same across roll-out strategies.

### 10. Vaccinating adolescents

We did not include those younger than 20 years of age in any baseline analysis as most vaccine products currently available are not authorised for such age groups in most countries in the WHO European Region at the time of this study. As results from clinical trials conducted among adolescents emerge, some vaccines are now authorised for use in those between 12 and 15 years of age in a small number of countries.[38] We thus expanded our analysis to include adolescents using the fastest vaccine roll-out scenario explored (i.e. R4) as it is the only one involving substantial vaccine surplus. We only expanded using V60 and V75 as the last groups vaccinated were younger adults, with whom it may make sense to potentially include adolescents. We found that vaccinating adolescents would bring additional health and economic benefits and that vaccinating adolescents simultaneously with younger adults was more beneficial than vaccinating them after the maximum uptake level among younger adults have been reached.

| Roll-out Strategy | Policy | Deaths | Cases | Adj. Life Expectancy | Quality Adj. Life Years | Human Capital |
|---|---|---|---|---|---|---|
| R4 | V60 | 9 | 20 | 12 | 11 | 10 |
| R4 | V60a | 17 | 5 | 10 | 16 | 18 |
| R4 | V60b | 12 | 13 | 10 | 11 | 10 |
| R4 | V75 | 11 | 13 | 10 | 11 | 11 |
| R4 | V75a | 27 | 25 | 28 | 27 | 27 |
| R4 | V75b | 11 | 13 | 10 | 11 | 11 |

**Table S8. Counts of cases where a policy is ranked first in terms of optimising health and economic benefits (including ties)**

**Caption:** The denominator for these proportions is 38, the number of countries within the WHO European Region without data availability or sparsity issues. In the expansion (a), adolescents were vaccinated with the last group in the baseline strategy; in the expansion (b), adolescents were vaccinated after the last group in the baseline strategy had reached their maximum uptake level.

### 11. Country-specific vaccine-prioritisation strategies by different vaccine profiles

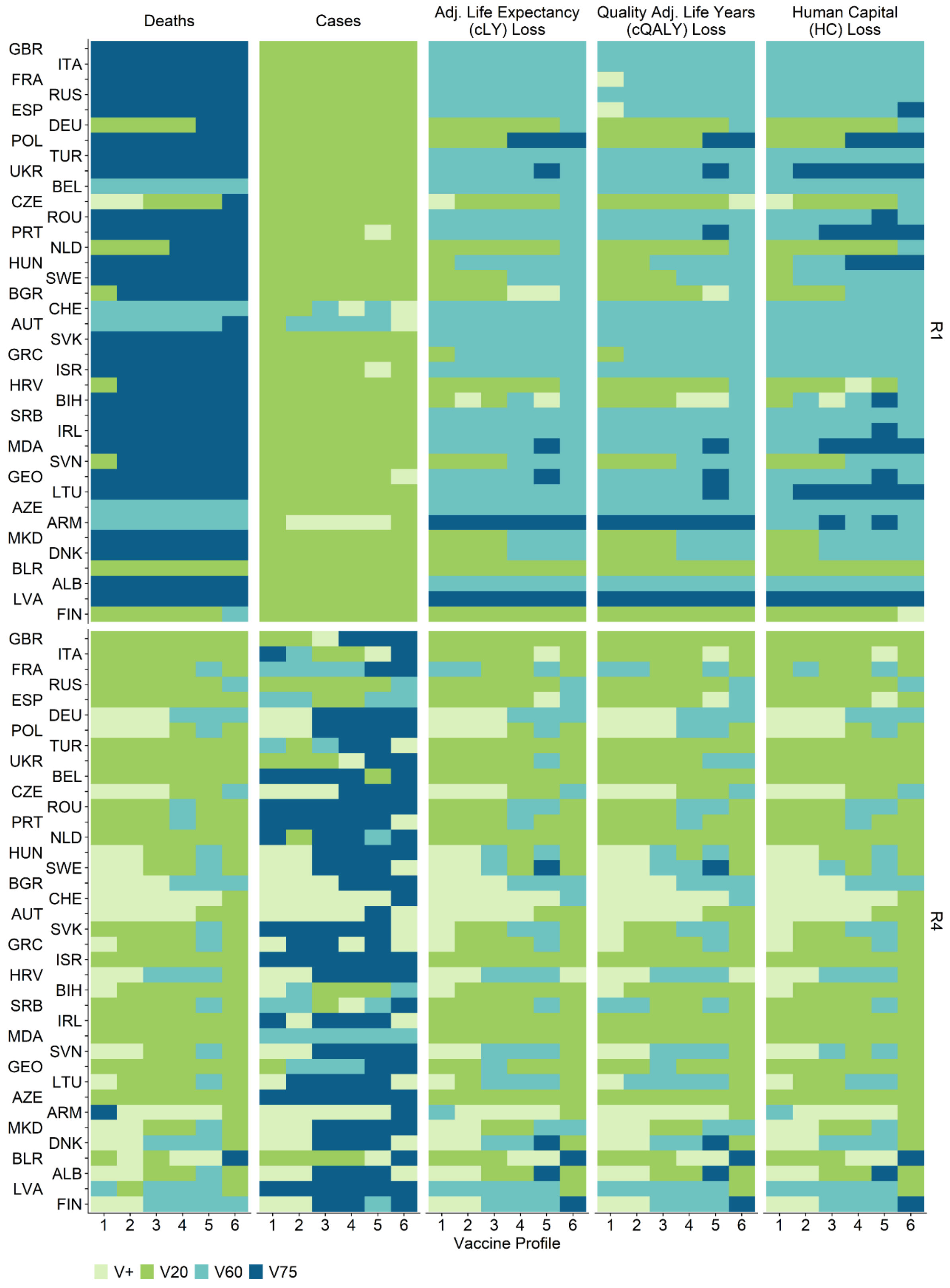

**Figure S16. Optimal vaccine prioritisation strategies for different vaccine characteristics under R1 and R4.**

**Caption:** Optimal strategy for each country and vaccine profile while minimising mortality, morbidity, adjusted life expectancy (cLE), quality-adjusted life-years (cQALY), or human capital (HC) losses for 38 countries in the WHO European Region with fitted models. Countries are arranged in the order of the expected proportion of the population no longer susceptible to SARS-CoV-2 on 01 Jan 2021 (descending).

23 Pfizer-BioNTech. Vaccines and Related Biological Products Advisory Committee Meeting December 10, 2020. US Food and Drug Administration, 2020.

24 World Health Organization. Global Health Observatory data repository | Life tables by WHO region - Europe. World Health Organization. https://apps.who.int/gho/data/view.main.LIFEEUR?lang=en (accessed April 21, 2021).

25 Briggs AH, Goldstein DA, Kirwin E, *et al.* Estimating (quality-adjusted) life-year losses associated with deaths: With application to COVID-19. *Health Economics* 2021; **30**: 699–707.

26 Szende A, Janssen B, Cabases J, editors. Self-Reported Population Health: An International Perspective based on EQ-5D. Dordrecht: Springer Netherlands, 2014 DOI:10.1007/978-94-007-7596-1.

27 World Health Organization. WHO guide for standardization of economic evaluations of immunisation programmes. https://apps.who.int/iris/bitstream/handle/10665/329389/WHO-IVB-19.10-eng.pdf (accessed April 21, 2021).

28 Hoek AJ van, Underwood A, Jit M, Miller E, Edmunds WJ. The Impact of Pandemic Influenza H1N1 on Health-Related Quality of Life: A Prospective Population-Based Study. *PLOS ONE* 2011; **6**: e17030.

29 Halpin SJ, McIvor C, Whyatt G, *et al.* Postdischarge symptoms and rehabilitation needs in survivors of COVID-19 infection: A cross-sectional evaluation. *Journal of Medical Virology* 2021; **93**: 1013–22.

30 Bennett S, Tafuro J, Mayer J, *et al.* Clinical features and outcomes of adults with coronavirus disease 2019: A systematic review and pooled analysis of the literature. *International Journal of Clinical Practice* 2021; **75**: e13725.

31 Cuthbertson BH, Roughton S, Jenkinson D, MacLennan G, Vale L. Quality of life in the five years after intensive care: a cohort study. *Crit Care* 2010; **14**: R6.

32 Griffiths J, Hatch RA, Bishop J, *et al.* An exploration of social and economic outcome and associated health-related quality of life after critical illness in general intensive care unit survivors: a 12-month follow-up study. *Crit Care* 2013; **17**: R100.

33 Greenhalgh T, Knight M, A'Court C, Buxton M, Husain L. Management of post-acute covid-19 in primary care. *BMJ* 2020; **370**: m3026.

34 The World Bank. GDP per capita, PPP (current international $). https://data.worldbank.org/indicator/NY.GDP.PCAP.PP.CD (accessed April 21, 2021).

35 Education, Audiovisual and Culture Executive Agency. The organisation of school time in Europe: primary and general secondary education: 2019/20. LU: Publications Office, 2019 https://data.europa.eu/doi/10.2797/678694 (accessed April 26, 2021).

36 US CDC. Public Health Guidance for Community-Related Exposure. Centers for Disease Control and Prevention. 2020; published online Feb 11. https://www.cdc.gov/coronavirus/2019-ncov/php/public-health-recommendations.html (accessed April 26, 2021).

37 Eurostat. GISCO data distribution API - Countries 2020. https://gisco-services.ec.europa.eu/distribution/v1/countries-2020.html (accessed April 12, 2021).

38 FDA Office of the Commissioner. FDA Authorizes Pfizer-BioNTech COVID-19 Vaccine for Emergency Use in Adolescents in Another Important Action in Fight Against Pandemic. FDA, 2021 https://www.fda.gov/news-events/press-announcements/coronavirus-covid-19-update-fda-authorizes-pfizer-biontech-covid-19-vaccine-emergency-use (accessed June 15, 2021).
